# Variant-specific symptoms of COVID-19 among 1,542,510 people in England

**DOI:** 10.1101/2022.05.21.22275368

**Authors:** Matthew Whitaker, Joshua Elliott, Barbara Bodinier, Wendy Barclay, Helen Ward, Graham Cooke, Christl A. Donnelly, Marc Chadeau-Hyam, Paul Elliott

**Author notes:** **Address for Correspondence:** Professor Paul Elliott, Department of Epidemiology and Biostatistics, School of Public Health, Imperial College London, St Mary’s campus, London W2 1PG, UK. Joint first author (Matthew Whitaker, Joshua Elliott). Equal contribution (Marc Chadeau-Hyam, Paul Elliott).

## Abstract

Infection with SARS-CoV-2 virus is associated with a wide range of symptoms. The REal-time Assessment of Community Transmission -1 (REACT-1) study has been monitoring the spread and clinical manifestation of SARS-CoV-2 among random samples of the population in England from 1 May 2020 to 31 March 2022. We show changing symptom profiles associated with the different variants over that period, with lower reporting of loss of sense of smell and taste for Omicron compared to previous variants, and higher reporting of cold-like and influenza-like symptoms, controlling for vaccination status. Contrary to the perception that recent variants have become successively milder, Omicron BA.2 was associated with reporting more symptoms, with greater disruption to daily activities, than BA.1. With restrictions lifted and routine testing limited in many countries, monitoring the changing symptom profiles associated with SARS-CoV-2 infection and induced changes in daily activities will become increasingly important.

---

A meta-analysis of studies from the first wave of the pandemic identified 30 symptoms reported in multiple studies,^1^ including common influenza-like symptoms (cough, fever, myalgia/fatigue, headache, sputum production), and less common but more specific symptoms including change or loss of sense of smell and taste.

Previous community-based studies have assessed the degree to which symptom data can predict polymerase chain reaction (PCR) positivity for SARS-CoV-2, and have used variable selection and ranking techniques to identify the most important (set of) symptoms for case identification.^2– 4^ Further studies have indicated that symptom profiles may differ between variants of SARS-CoV-2.^5–7^

The relationship between symptom profile and cycle threshold (Ct) value from PCR testing (an established proxy for viral load,^8–10^ which in turn correlates with infectiousness^11,12^) has also yet to be fully investigated. Identifying individuals who are more likely to be i) infected, and ii) infectious on the basis of symptom profile would have clinical value as governments move away from mass testing programmes and mandatory isolation measures.

Here, we use regression modelling and variable selection models in the large community-based REal-time Assessment of Community Transmission -1 (REACT-1) study that was in the field approximately monthly from 1 May 2020 to 31 March 2022 to i) describe the symptom profiles of the main variants of SARS-CoV-2 that have been dominant in England and worldwide since May 2020, namely wild-type, Alpha, Delta and Omicron BA.1 and BA.2, and ii) identify the symptoms that are most predictive of high viral load, and hence infectiousness, for each variant.

## Methods

### Study population

The REACT-1 study has been tracking the prevalence of SARS-CoV-2 in the general population of England. The study protocol and methodology have been published in detail;^2,13^ briefly, every 4–6 weeks, recruitment letters were sent to a random, nationally representative sample of people aged 5 years and over in England, using the National Health Service patient register. Participants then obtained self-administered throat and nasal swabs for SARS-CoV-2 PCR testing and completed an online or telephone questionnaire which included questions on demographic variables, behaviour, and recent symptoms. Questionnaires for each of the 19 completed rounds since May 2020 are available on the study website (https://www.imperial.ac.uk/medicine/research-and-impact/groups/react-study/for-researchers/react-1-study-materials/). Between 95,000 and 175,000 viable swabs and valid responses were gathered each round, with respondents unaware of their test result at the time of their response.

Participants were asked whether they experienced any of a list of 26 potential COVID-19 symptoms in the week prior to their test. These included loss or change of sense of smell or taste, respiratory/cardiac symptoms (new persistent cough, chest pain, tight chest, shortness of breath), cold-like symptoms (runny nose, blocked nose, sneezing, sore throat, hoarse voice, sore eyes), influenza-like symptoms (fever, chills, muscle aches, headache), gastrointestinal symptoms (nausea/vomiting, abdominal pain/belly ache, diarrhoea, appetite loss), fatigue-related symptoms (tiredness, severe fatigue, difficulty sleeping), and others (dizziness, heavy arms or legs, numbness/tingling).

We use data from 15 rounds of REACT-1 between 19 June 2020 and 31 March 2022 into distinct phases that correspond with the dominance of different SARS-CoV-2 variants in England: rounds 2–7 (at approximately monthly intervals between 19 June and 3 December 2020), when wild-type was dominant; rounds 8–10 (between 6 January and 29 March 2021), when Alpha (B.1.1.7) was dominant; rounds 13–15 (between 24 June and 5 November 2021), when Delta (B.1.617.2) was dominant; and rounds 17–19 (between 5 January and 31 March 2022), when Omicron (B.1.1.529) was dominant. In rounds 17–19 we use sequencing data to identify those participants who were infected with BA.1 or BA.2. Round 1 is excluded because the symptom questions asked were not consistent with subsequent rounds. Rounds 11, 12 and 16 are excluded from analysis because they occurred at times when two variants were competing for dominance in the population.^14^

Adults aged 18 years and over were included in the analysis. A total of 266,847 participants were excluded because of missing symptom data, and 38 were excluded because of missing age or sex data resulting in a final study population, after exclusions, of 1,542,510 participants.

### Statistical analyses

We used univariable logistic regression models to estimate the risk of PCR swab-positivity for each variant conditional on experiencing each of the 26 symptoms. Models were adjusted on age group, sex, and self-reported vaccination status (coded as the number of vaccines received). Odds ratios and 95% confidence intervals are reported for each symptom and each variant.

Variable selection models were trained on 70% of the data set, with 30% held back for model performance evaluation (see **Supplementary Methods**). We used stability selection applied to least absolute shrinkage and selection operator (LASSO) penalised logistic regression, with swab positivity as the binary outcome variable, and the 26 symptoms as predictors. To adjust for age, sex and vaccination status, these were included in the models as unpenalised variables. The regression coefficients for selected symptoms were constrained to non-negative values. LASSO models were fit on 1000 random 50% subsamples of the 70% training data. The proportion of models in which each symptom was selected is taken as a measure of variable importance. The threshold in selection proportion for final variable selection was calibrated in conjunction with the LASSO penalty parameter using an internal stability score.^15^

#### BA.1 vs BA.2

Omicron BA.1 and BA.2 lineages were determined using viral genome sequencing on swab-positive swabs from rounds 17–19. We compared the symptom profiles in (BA.1 or BA.2) swab-positive individuals using logistic regression with BA.2 vs BA.1 as the binary outcome variable and each of the 26 symptoms as explanatory variables, adjusted on age group, sex, vaccination status and round. As a sensitivity analysis, we 1:1 matched swab-positive participants with BA.1 and BA.2 on age group (+/-5 years), sex, vaccination status and round in rounds 17–19, and conducted conditional logistic regression with BA.2 vs BA.1 as the binary outcome variable.

#### Severity of symptoms

To assess whether there are differences in symptom severity between BA.1 and BA.2 independent of vaccination history we took a subset of swab-positive individuals with sequence-confirmed BA.1 or BA.2 who had received booster (third) vaccines at least two weeks before their PCR test. In this group, we compared the risk of reporting symptoms that affected their daily activities ‘a lot’ vs ‘a little’ or ‘not at all’ in people infected with BA.1 vs BA.2.

We adjusted for age, sex, days since booster (including a squared polynomial term), and round.

#### Ct values

Finally, we investigated the relationship between N-gene Ct value and symptom profile among swab positive individuals in rounds 17–19 (>95% Omicron), using linear regression models with Ct value as the outcome variable and each symptom separately as the independent variable. We also compared Ct values between swab-positive individuals with BA.1 and BA.2 using an unpaired Wilcoxon test.

## Results

### Descriptive and univariable analysis

The characteristics of our study population are summarised in Figure 1 and Supplementary Tables 1 and 2. We included a total of 17,448 swab positive individuals: 2,971 (0.4% [0.4,0.4] unweighted prevalence) for wild,type; 2,275 (0.6% [0.6,0.7]) for Alpha; 1,493 (0.7% [0.6,0.7]) for Delta and 10,709 (4.4% [4.3,4.5]) for Omicron (Table S1).

**Figure 1.**
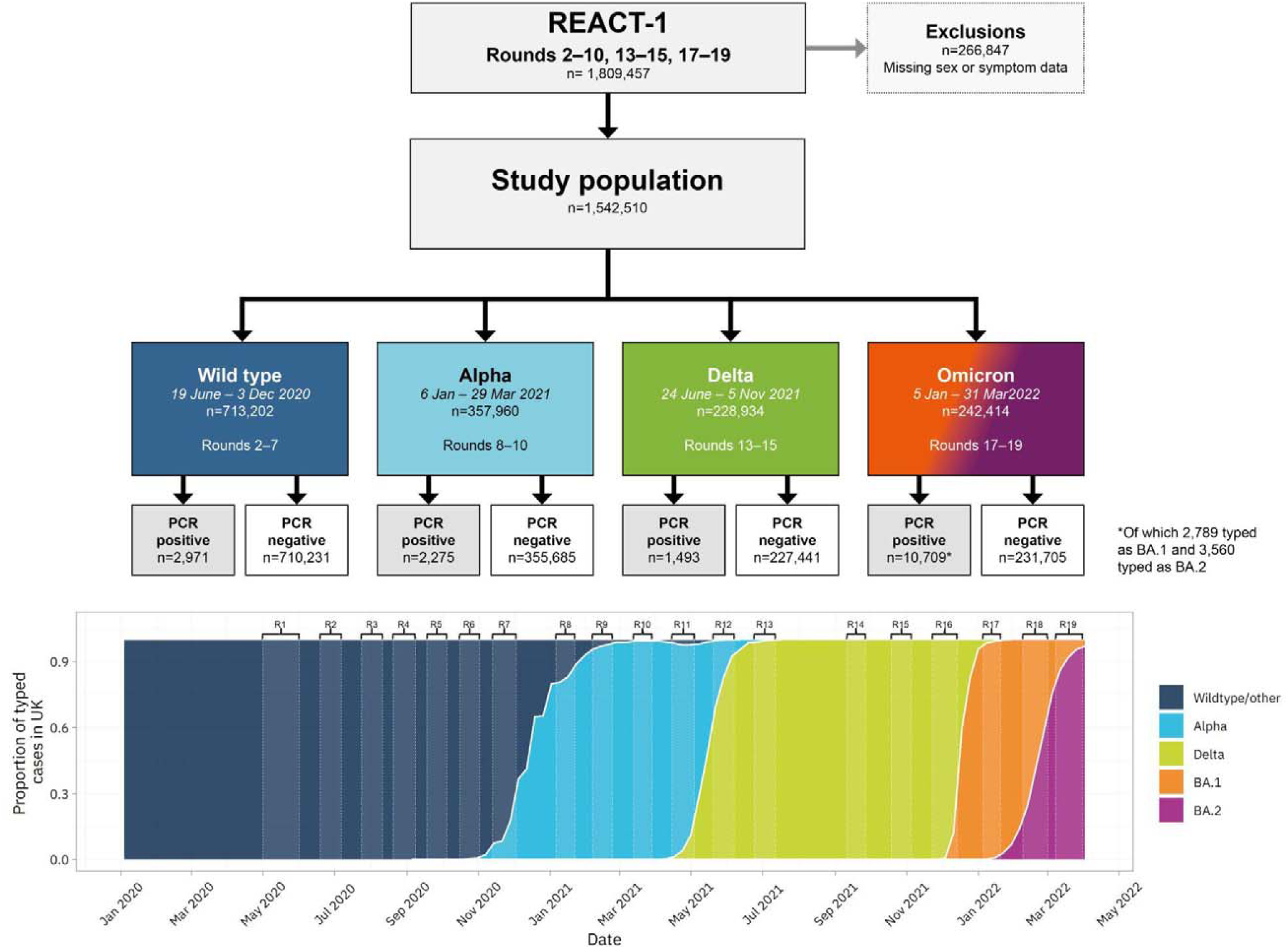
Study population flow-chart. Variant prevalence data in bottom panel is from GISAID^23^

The proportion of swab positive individuals reporting any of 26 symptoms was highest in those infected with BA.2 (75.9% [74.4,77.2], compared with 70% [68.3,71.6] in those with BA.1, 63.8% [61.3,66.2] in those with Delta, 54.7%, [52.7,56.8] in those with Alpha and 45% [43.3,46.8] in those with wild-type) (Table S2). Background prevalence of symptoms was also highest during January – March 2022, when Omicron dominated: 21.9%, [21.7,22.0] of all respondents reported one or more symptoms, compared with 13.5% [13.4,13.5] during the wild-type period (Table S1).

Those infected with BA.2 reported an average of 6.02 symptoms in the week prior to PCR testing, compared with 2.70, 3.38, 4.63 and 4.63 for wild-type, Alpha, Delta and BA.1 respectively (Table S2). A larger proportion of people with BA.2 reported that their symptoms had affected their ability to carry out day-to-day activities ‘a lot’ (17.6% [16.3,18.8]) compared with those infected with BA.1 (10.7% [9.6,11.9]) or Delta (10.5%, [9.1,12.2]) (Table S2).

All symptoms were positively associated with swab positivity for all variants (Figure 2, Table S3). The odds ratio for swab positivity of ‘any of 26 symptoms’ was highest for BA.2 (OR=12.9 [11.9,14.0], compared with 5.16 [4.79,5.55], 6.01 [5.12,7.06], 9.53 [8.55,10.6] and 9.61 [8.82,10.5] for wild-type, Alpha, Delta and BA.1, respectively) (Table S3, Figure 2).

**Figure 2.**
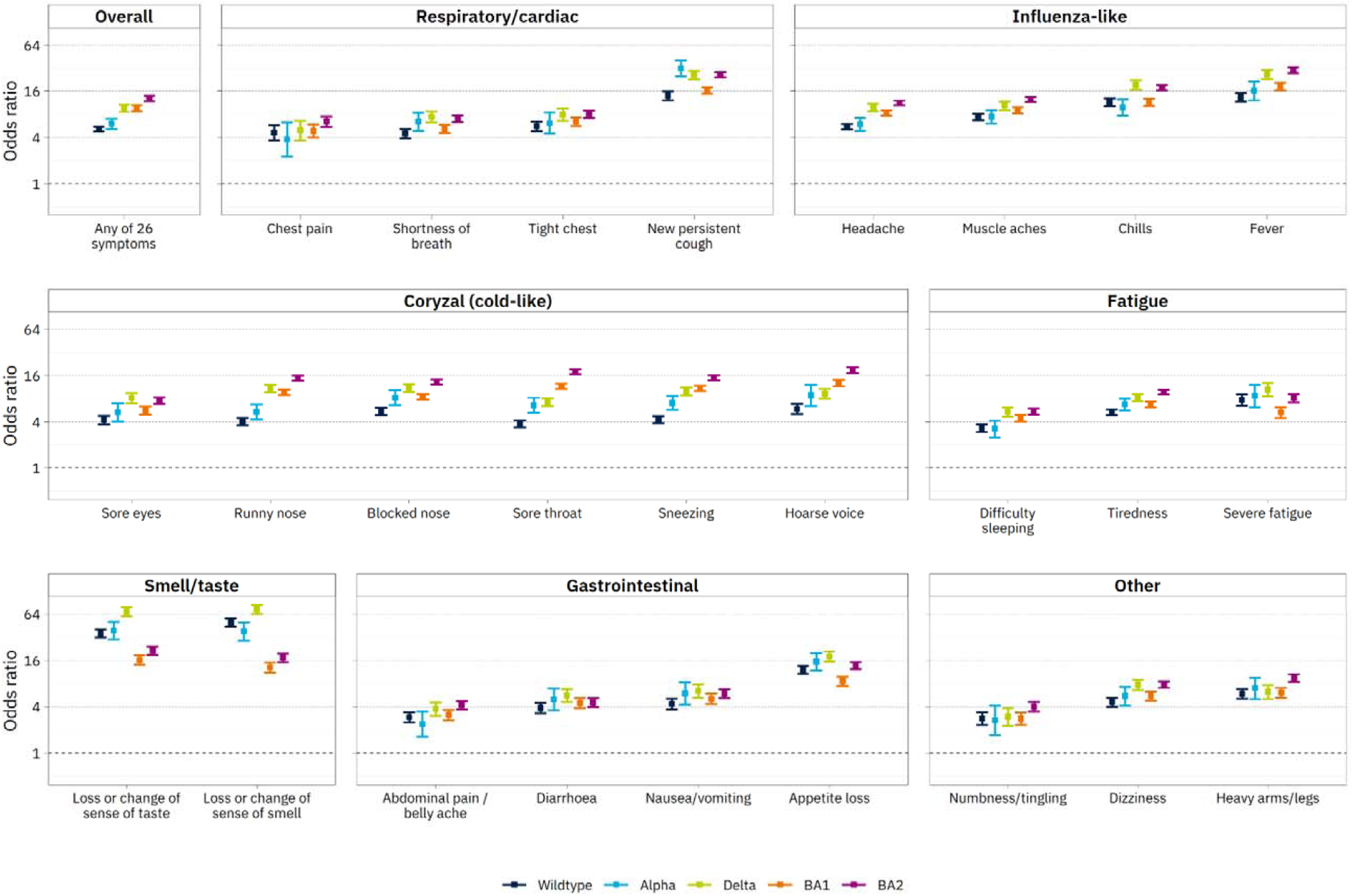
Comparison of ORs for swab positivity based on presence or absence of any of 26 symptoms surveyed across five variant-phases of REACT-1. ORs are derived from logistic regression models with swab positive (1/0) as the outcome variable, adjusted on age, sex and vaccination status. Bars show 95% confidence intervals. ORs are higher in BA.2 than BA.1 for all symptoms. Fever and cough have the highest ORs for BA.1 and BA.2, while loss of smell and taste have the highest ORs in all previous variants.

Unlike for wild-type, Alpha, and Delta, where the highest odds ratios for swab positivity were for loss or change of sense of smell (ORs 49.7 [44.3,55.7], 37.8 [28.6,50.0] and 73.4 [64.2,83.9] respectively) or taste (ORs 35.9 [31.9,40.4], 38.9 [29.9,50.6] and 68.1 [59.4,78.0] respectively), for BA.1 and BA.2, influenza-like and cold-like symptoms were relatively more predictive of swab positivity, and loss or change of sense of smell or taste relatively less so. Within BA.1 and BA.2, the highest odds ratio of all symptoms was for fever: ORs were 18.4 [16.5,20.5] in BA.1 and 30.2 [27.7,33.0] in BA.2, compared with 12.9 [11.1,15.1] and 17.2 [15.1,19.5] respectively for loss or change of sense of smell and 16.0 [13.9,18.5] and 21.3 [18.9,24.0] respectively for loss or change of sense of taste (Figure 2, Table S3).

### Multivariable analysis for variable selection

We used LASSO penalised logistic regression to identify parsimonious symptom sets selected as jointly and positively predictive of swab positivity for each variant (Figure 3); this method takes into account differences in symptom cooccurrence by variant (Figure S3). Loss or change of sense of taste, new persistent cough, and fever were selected for each variant. Notably, cold-like symptoms of runny nose, sore throat, sneezing and hoarse voice were only selected for Omicron (BA.1 and BA.2).

**Figure 3.**
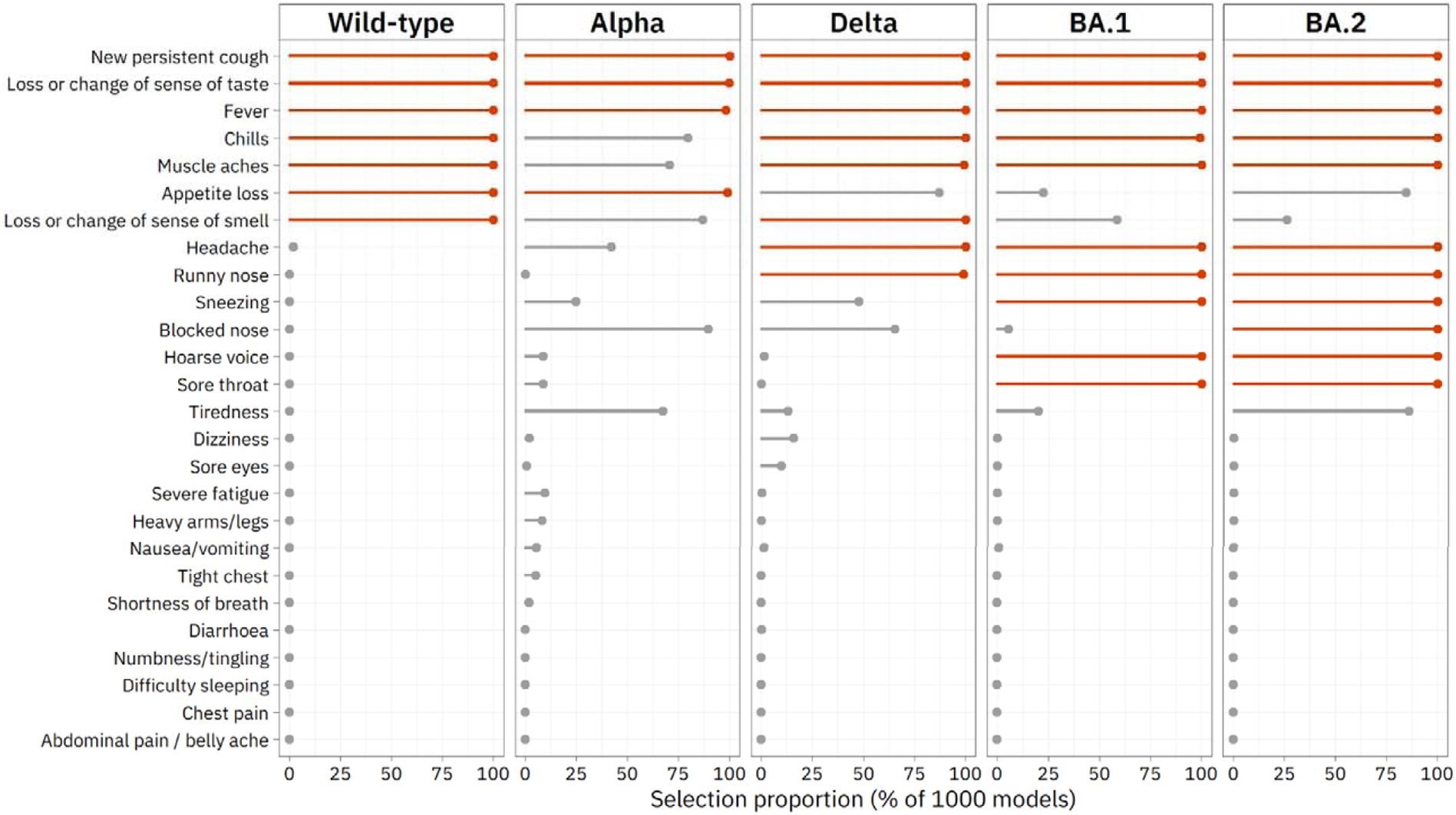
Results of LASSO stability selection proportions with swab positive/negative as the binary outcome variable and each of 26 symptoms as predictors, in five SARS-CoV-2 variants in England. Age, sex and, where appropriate, vaccination status are forced into the models as unpenalised variables; regression coefficients for the symptoms are constrained to be positive. The selection proportion indicates the proportion of LASSO models, trained on subsamples of the data, in which each symptom was selected as a predictor.

### Omicron (BA.1 and BA.2)

Comparing symptoms for BA.2 vs BA.1 using (adjusted and matched) logistic regression (see **Methods**), infection with BA.2 was positively associated with chest pain, severe fatigue, runny nose, muscle aches, sneezing, fever, chills, tiredness, blocked nose and headache (in both models); in unmatched analysis, infection with BA.2 is further associated with further associated with sore eyes, appetite loss and new persistent cough.

In a triple-vaccinated subgroup of 4,834 swab-positive individuals with BA.1 or BA.2, those infected with BA.2 were 64% more likely to report symptoms that interfered with their ability to carry out day-to-day activities ‘a lot’ (OR 1.64 [1.22, 2.19]) vs ‘a little’, ‘not at all’, or not reporting any symptoms, after adjustment for age group, sex, round, and time since third vaccine (Table 1). In the same models, men were 40% less likely than women to report symptoms that interfered with their ability to carry out day-to-day activities ‘a lot’ (OR 0.60, [0.51,0.72]).

**Table 1.** Results from logistic regression modelling of the response to the question “How much, if at all, do the symptoms your/your child have/has had in the last 7 days reduce your/their ability to carry out day-to-day activities?” as a function of BA.1 / BA.2 infection, age, sex group, days since booster, and round, among 4,834 triple-vaccinated swab-positive individuals with either BA.1 or BA.2 lineage. The outcome variable is a binary 1/0 indicating where 1 indicates that the individual responded “A lot” and 0 indicates that the individual responded “A little” or “Not at all”, or reported that they were asymptomatic. Those who responded “Prefer not to say” or “Don’t know” were excluded from modelling.

| Independent variable | Odds ratio with CI | P-value |
| --- | --- | --- |
| Intercept | 0.04, [0.02,0.07] | <0.01* |
| Age group: 25-34 | 1.70, [0.98,2.94] | 0.06 |
| Age group: 35-44 | 2.39, [1.41,4.07] | <0.01* |
| Age group: 45-54 | 2.91, [1.71,4.96] | <0.01* |
| Age group: 55-64 | 3.11, [1.83,5.28] | <0.01* |
| Age group: 65-74 | 1.75, [1.01,3.04] | 0.05* |
| Age group: 74+ | 1.89, [1.02,3.49] | 0.04* |
| Sex: male | 0.60, [0.51,0.72] | <0.01* |
| **BA.2** | **1.64, [1.24,2.19]** | **<0.01\*** |
| Days since booster vaccine | 1.00, [1.00,1.01] | 0.13 |
| [Days since booster vaccine]^2 | 1.00, [1.00,1.01] | 0.22 |
| Round 18 | 1.26, [0.90,1.75] | 0.18 |
| Round 19 | 1.23, [0.81,1.86] | 0.33 |

#### Ct values

Ct values were lower for BA.2 than BA.1 (Figure S1). This may reflect the timing of the sampling with respect to the growth of the variant since more recent infections will tend to have lower Ct values (see Table S2 and Supplementary Figure S5, which show that mean time since symptom onset was lower in BA.2 than for BA.1). Symptomatic individuals had lower Ct values than asymptomatic people. In linear regression models among swab positive individuals in rounds 17–19, for each of the 26 surveyed symptoms, symptom reporting was associated with a lower Ct value. The lowest adjusted Ct values were for influenza-like or cold-like symptoms: fever, chills, sore throat, muscle aches, runny nose, sneezing and headache (Figure 4), which frequently co-occurred (Figure S3). With the exception of fever, these symptoms were also commonly reported as the first symptom among symptomatic swab positives (Figure S4, Table S2).

**Figure 4.**
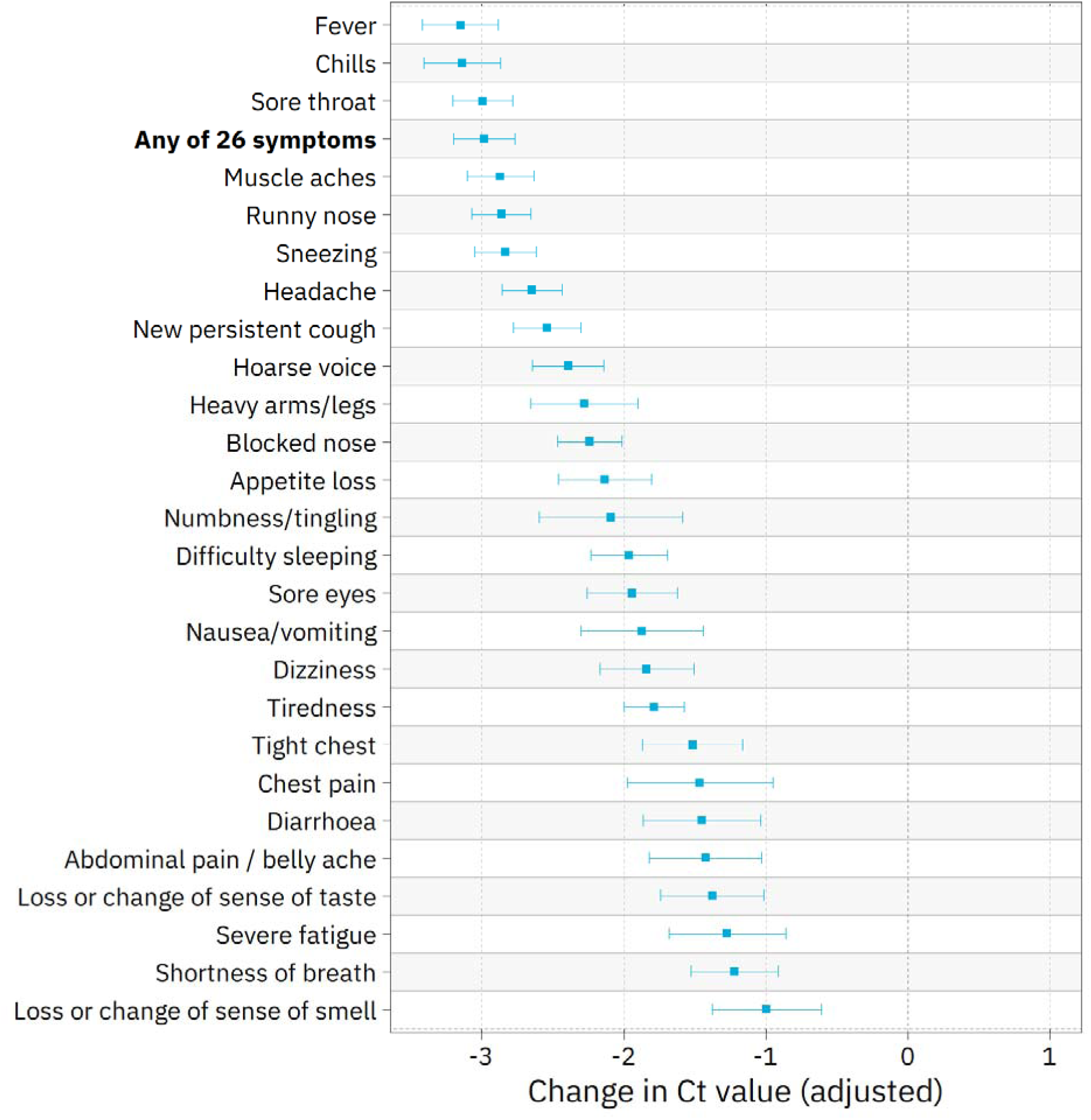
Results of linear regression models with N-gene Ct values as the outcome variable and symptoms as individual predictors, adjusted for age, sex and, where appropriate, vaccination status, among swab-positive respondents in rounds 17–19. Fever, chills, and sore throat are the symptoms with the strongest negative association with Ct value, each associated with approximately a tenfold decrease in viral load (3 Ct ∼ 10x Δ).

**Figure 5.**
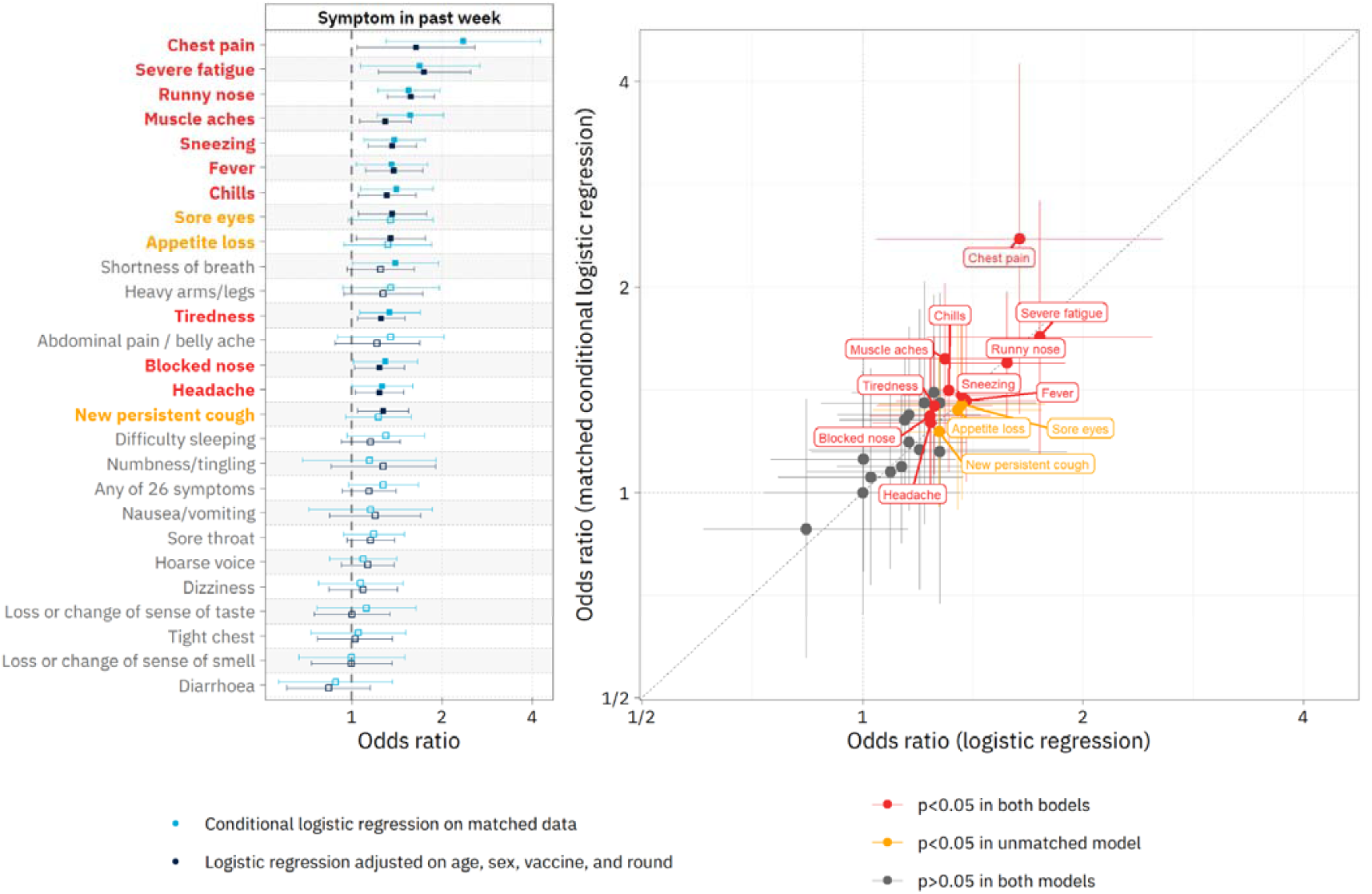
ORs for infection with BA.2 vs BA.1 among swab-positive respondents. ORs are derived from (i) logistic regression models with BA.2 vs BA.1 as the binary outcome variable, and presence or absence of any of 26 symptoms as explanatory variables, adjusted on age group, sex, round and vaccination status, among 5,598 swab-positive individuals with either BA.2 or BA.1 in rounds 17–19; and (ii) conditional logistic regression models with BA.2 vs BA.1 as the outcome variable among 1,510 swab-positive individuals with either the BA.2 or BA.1 variant in rounds 17–19, matched 1:1 on age (+/-5 years), sex, vacination status and round. In left panel, bars show 95% confidence intervals, and symptoms are ordered by mean OR across both models. Right panel directly plots the ORs from the two models for comparison. In both analyses, infection with BA.2 (vs BA.1) is positively associated with chest pain, severe fatigue, runny nose, muscle aches, sneezing, fever, chills, tiredness, blocked nose and headache; in unmatched analysis, infection with BA.2 is further associated with sore eyes, appetite loss and new persistent cough.

## Discussion

In this study of more than 1.5 million people randomly selected from the population in England, we show a change in symptom reporting associated with Omicron compared with previous variants, and within Omicron for BA.2 vs BA.1. This may reflect changes in the underlying pathophysiology associated with different variants, affecting receptor binding, cell entry and host response, against a background of varying levels of population immunity (both from natural infection and vaccine-induced).^16–18^

We found that loss or change of sense of smell and taste were less predictive of swab positivity for Omicron than for other variants, and that cold-like symptoms were more predictive for Omicron than for previous variants. Both these findings were consistent with previous research.^5,19,20^ It is of interest that infections with Omicron variants are not as strongly associated with anosmia. The loss of sense of smell and taste following infection with early variants of SARS-CoV-2 results from the downregulated expression of olfactory receptors.^21^ It is likely that changes in the sequence of viral genes that regulate host responses in Omicron do not result in this effect. Detailed transcriptomic studies in animal models and humans may help to pinpoint the mechanisms involved.

Comparing Omicron BA.2 with BA.1, we found that those with BA.2 were more likely to be symptomatic, reported more symptoms on average, were more likely to report a number of influenza-like and cold-like symptoms, and were more likely to report that their symptoms affected their day-to-day activities ‘a lot’. This last finding was robust to adjustment for time since third vaccine dose and is therefore unlikely to be explained by waning immunity from vaccination.

From 1 April 2022 the UK government moved to a policy of ‘living with COVID’.^22^ With the lifting of restrictions and access to free testing limited, identifying individuals who are particularly likely to be infectious on the basis of symptoms alone may help reduce ongoing transmission of SARS-CoV-2. We showed that reporting fever, chills, sore throat, muscle aches, runny nose, sneezing and headache was associated with the lowest adjusted Ct values and therefore most likely to be indicative of higher viral load and increased infectiousness.

Our study has limitations. Response rates varied between 11.7% and 26.5% for rounds 2–19, so the samples may not be fully representative of, or results fully generalisible to, the population. Nevertheless, our random community sampling procedure included individuals from all of the 315 lower tier local authority areas in England in each round, ensuring wide geographical coverage and socio-economic and demographic diversity. Of those who provided valid swabs and consented to linkage in rounds 1–19 of REACT-1 (2,191,597 people in total), approximately 3% (65,915 people) participated in more than one round. On this basis, a correction factor of 1.015 could therefore be applied to the standard error estimates. We are not able to definitively identify instances of participation in more than one round among those who did not consent to linkage. However, because the consent-based estimate of the correction factor is so close to one, we feel confident reporting uncorrected SEs and confidence intervals. The symptoms surveyed were not exhaustive but, while not specific to COVID-19, were all shown to be predictive of SARS-CoV-2 swab positivity. Our analysis covers a period of 22 months, during which time background levels of natural and vaccine-acquired immunity varied substantially, making it difficult to differentiate the effect of viral mutations from the impact of vaccines and prior infection.^18^ As REACT-1 data collection was non-continuous, we may have captured different stages of epidemic growth across variants, which may have differentially affected symptom reporting at different times.

In summary, we have detected changes in symptom profiles reported during nearly two years of the epidemic in England, reflecting the emergence of different variants over that period. Most recently, infection with Omicron is associated with lower reporting of loss or change of sense of smell and taste, and higher reporting of cold-like and influenza-like symptoms. Sequence-confirmed BA.2 was associated with reporting of more symptoms and greater disruption to daily activity compared with BA.1. As routine testing becomes more limited in many countries, and as new variants emerge, understanding the symptom profiles which can identify individuals with a higher risk of transmission will become increasingly important.

## Data Availability

Access to REACT-1 individual-level data is restricted to protect participants' anonymity.
Summary statistics, descriptive tables, and code from the current REACT-1 study are available at https://github.com/mrc-ide/reactidd (doi 10.5281/zenodo.6550327). REACT-1 study materials are available for each round at
https://www.imperial.ac.uk/medicine/research-and-impact/groups/react-study/react-1-study-materials/
Sequence read data are available without restriction from the European Nucleotide Archive at https://www.ebi.ac.uk/ena/browser/view/PRJEB37886, and consensus genome sequences are available from the Global initiative on sharing all influenza data (GISAID).

https://github.com/mrc-ide/reactidd/tree/master/inst/extdata/

## Ethics

We obtained research ethics approval from the South Central-Berkshire B Research Ethics Committee (IRAS ID: 283787). Notification of favorable opinion and brief summary of the protocol are available here: https://www.hra.nhs.uk/planning-and-improving-research/application-summaries/research-summaries/react1-covid-19-uph/

## Public involvement

A Public Advisory Panel provides input into the design, conduct, and dissemination of the REACT research program.

## Data availability

Access to REACT-1 individual-level data is restricted to protect participants’ anonymity. Summary statistics, descriptive tables, and code from the current REACT-1 study are available at https://github.com/mrc-ide/reactidd (doi 10.5281/zenodo.6550327). REACT-1 study materials are available for each round at https://www.imperial.ac.uk/medicine/research-and-impact/groups/react-study/react-1-study-materials/Sequence read data are available without restriction from the European Nucleotide Archive at https://www.ebi.ac.uk/ena/browser/view/PRJEB37886, and consensus genome sequences are available from the Global initiative on sharing all influenza data (GISAID)^23^.

## Supplementary material

**Table S1.**
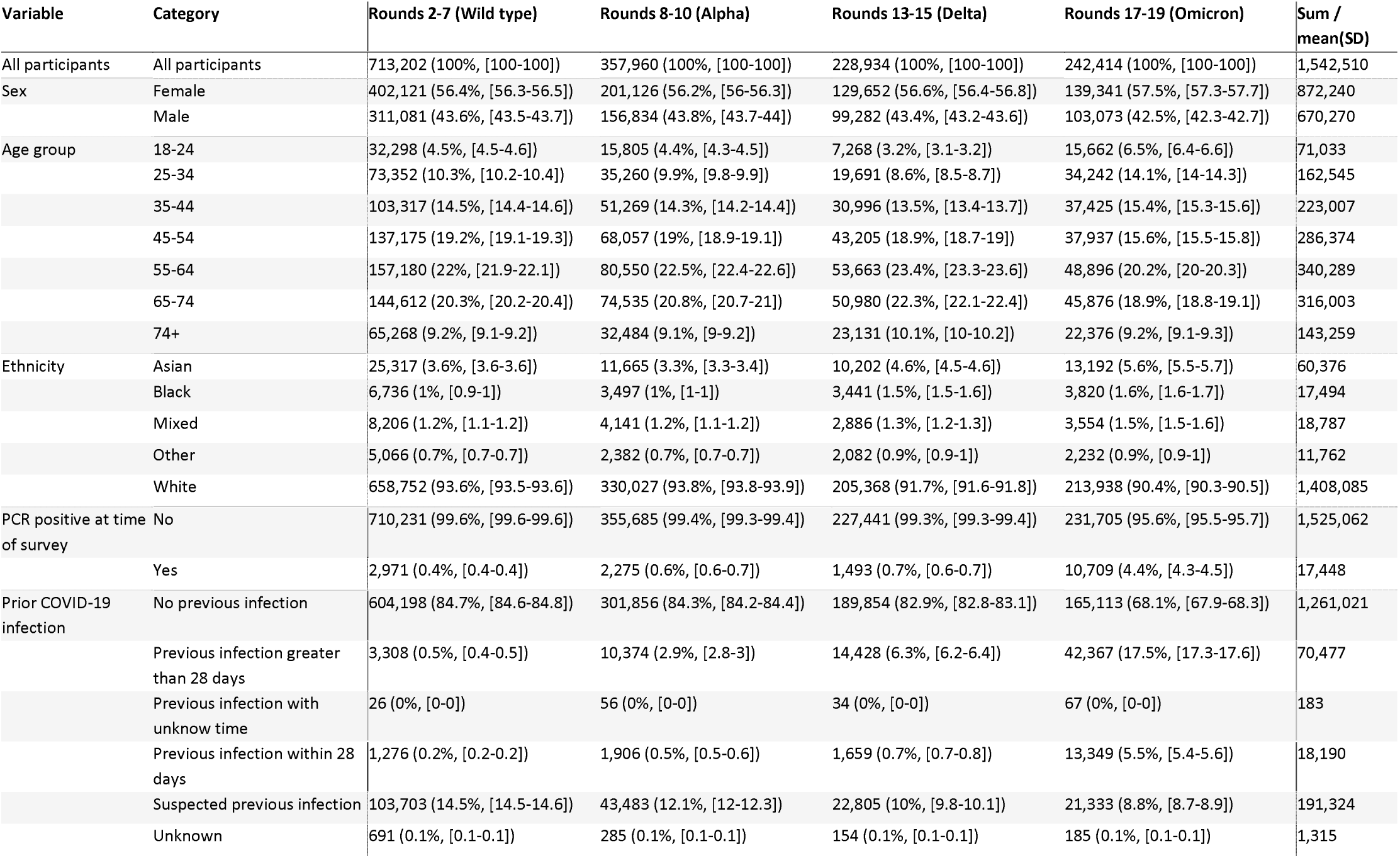
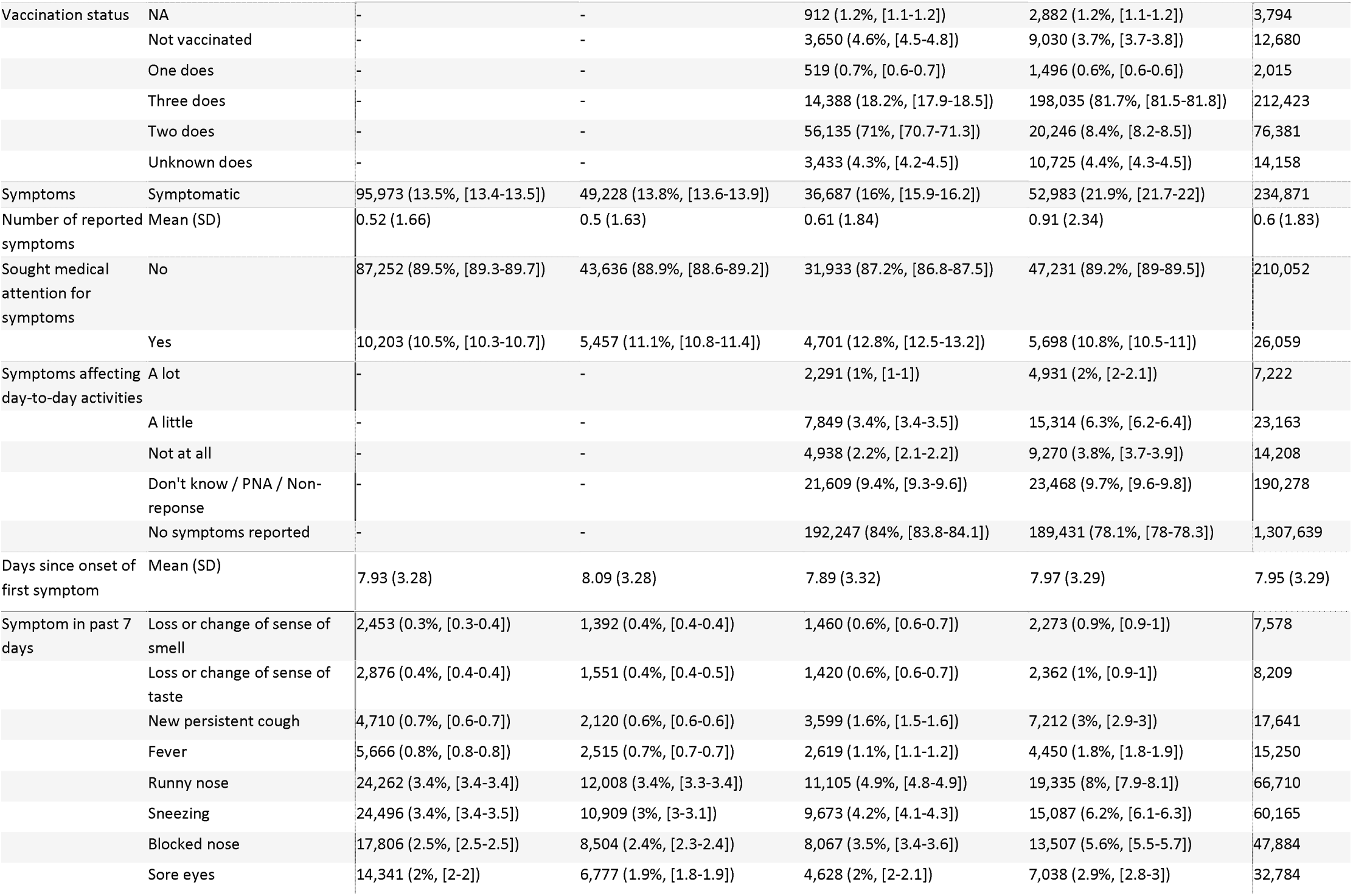
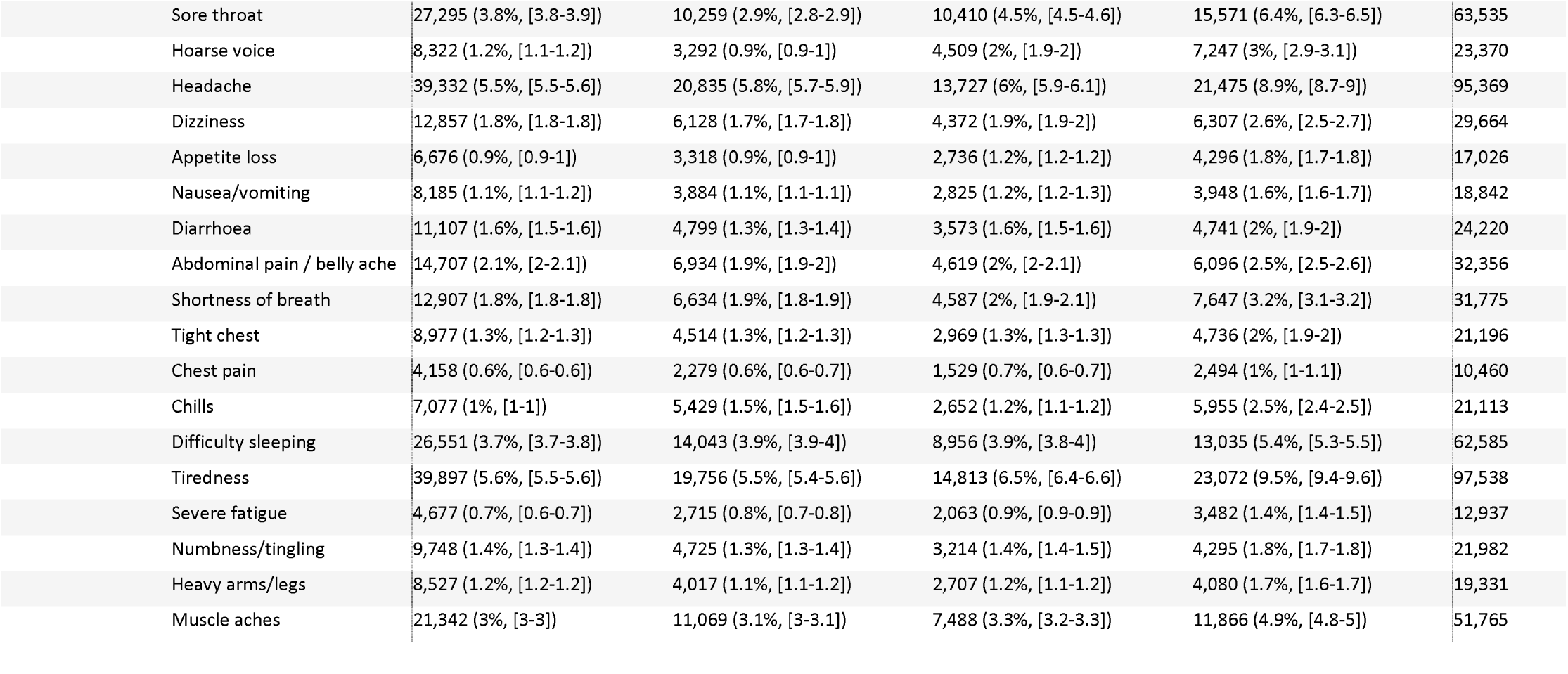
Characteristics of study population

| Variable | Category | Rounds 2-7 (Wild type) | Rounds 8-10 (Alpha) | Rounds 13-15 (Delta) | Rounds 17-19 (Omicron) | Sum / mean(SD) |
| --- | --- | --- | --- | --- | --- | --- |
| All participants | All participants | 713,202 (100%, [100-100]) | 357,960 (100%, [100-100]) | 228,934 (100%, [100-100]) | 242,414 (100%, [100-100]) | 1,542,510 |
| Sex | Female | 402,121 (56.4%, [56.3-56.5]) | 201,126 (56.2%, [56-56.3]) | 129,652 (56.6%, [56.4-56.8]) | 139,341 (57.5%, [57.3-57.7]) | 872,240 |
|  | Male | 311,081 (43.6%, [43.5-43.7]) | 156,834 (43.8%, [43.7-44]) | 99,282 (43.4%, [43.2-43.6]) | 103,073 (42.5%, [42.3-42.7]) | 670,270 |
| Age group | 18-24 | 32,298 (4.5%, [4.5-4.6]) | 15,805 (4.4%, [4.3-4.5]) | 7,268 (3.2%, [3.1-3.2]) | 15,662 (6.5%, [6.4-6.6]) | 71,033 |
|  | 25-34 | 73,352 (10.3%, [10.2-10.4]) | 35,260 (9.9%, [9.8-9.9]) | 19,691 (8.6%, [8.5-8.7]) | 34,242 (14.1%, [14-14.3]) | 162,545 |
|  | 35-44 | 103,317 (14.5%, [14.4-14.6]) | 51,269 (14.3%, [14.2-14.4]) | 30,996 (13.5%, [13.4-13.7]) | 37,425 (15.4%, [15.3-15.6]) | 223,007 |
|  | 45-54 | 137,175 (19.2%, [19.1-19.3]) | 68,057 (19%, [18.9-19.1]) | 43,205 (18.9%, [18.7-19]) | 37,937 (15.6%, [15.5-15.8]) | 286,374 |
|  | 55-64 | 157,180 (22%, [21.9-22.1]) | 80,550 (22.5%, [22.4-22.6]) | 53,663 (23.4%, [23.3-23.6]) | 48,896 (20.2%, [20-20.3]) | 340,289 |
|  | 65-74 | 144,612 (20.3%, [20.2-20.4]) | 74,535 (20.8%, [20.7-21]) | 50,980 (22.3%, [22.1-22.4]) | 45,876 (18.9%, [18.8-19.1]) | 316,003 |
|  | 74+ | 65,268 (9.2%, [9.1-9.2]) | 32,484 (9.1%, [9-9.2]) | 23,131 (10.1%, [10-10.2]) | 22,376 (9.2%, [9.1-9.3]) | 143,259 |
| Ethnicity | Asian | 25,317 (3.6%, [3.6-3.6]) | 11,665 (3.3%, [3.3-3.4]) | 10,202 (4.6%, [4.5-4.6]) | 13,192 (5.6%, [5.5-5.7]) | 60,376 |
|  | Black | 6,736 (1%, [0.9-1]) | 3,497 (1%, [1-1]) | 3,441 (1.5%, [1.5-1.6]) | 3,820 (1.6%, [1.6-1.7]) | 17,494 |
|  | Mixed | 8,206 (1.2%, [1.1-1.2]) | 4,141 (1.2%, [1.1-1.2]) | 2,886 (1.3%, [1.2-1.3]) | 3,554 (1.5%, [1.5-1.6]) | 18,787 |
|  | Other | 5,066 (0.7%, [0.7-0.7]) | 2,382 (0.7%, [0.7-0.7]) | 2,082 (0.9%, [0.9-1]) | 2,232 (0.9%, [0.9-1]) | 11,762 |
|  | White | 658,752 (93.6%, [93.5-93.6]) | 330,027 (93.8%, [93.8-93.9]) | 205,368 (91.7%, [91.6-91.8]) | 213,938 (90.4%, [90.3-90.5]) | 1,408,085 |
| PCR positive at time of survey | No | 710,231 (99.6%, [99.6-99.6]) | 355,685 (99.4%, [99.3-99.4]) | 227,441 (99.3%, [99.3-99.4]) | 231,705 (95.6%, [95.5-95.7]) | 1,525,062 |
|  | Yes | 2,971 (0.4%, [0.4-0.4]) | 2,275 (0.6%, [0.6-0.7]) | 1,493 (0.7%, [0.6-0.7]) | 10,709 (4.4%, [4.3-4.5]) | 17,448 |
| Prior COVID-19 infection | No previous infection | 604,198 (84.7%, [84.6-84.8]) | 301,856 (84.3%, [84.2-84.4]) | 189,854 (82.9%, [82.8-83.1]) | 165,113 (68.1%, [67.9-68.3]) | 1,261,021 |
|  | Previous infection greater than 28 days | 3,308 (0.5%, [0.4-0.5]) | 10,374 (2.9%, [2.8-3]) | 14,428 (6.3%, [6.2-6.4]) | 42,367 (17.5%, [17.3-17.6]) | 70,477 |
|  | Previous infection with unknow time | 26 (0%, [0-0]) | 56 (0%, [0-0]) | 34 (0%, [0-0]) | 67 (0%, [0-0]) | 183 |
|  | Previous infection within 28 days | 1,276 (0.2%, [0.2-0.2]) | 1,906 (0.5%, [0.5-0.6]) | 1,659 (0.7%, [0.7-0.8]) | 13,349 (5.5%, [5.4-5.6]) | 18,190 |
|  | Suspected previous infection | 103,703 (14.5%, [14.5-14.6]) | 43,483 (12.1%, [12-12.3]) | 22,805 (10%, [9.8-10.1]) | 21,333 (8.8%, [8.7-8.9]) | 191,324 |
|  | Unknown | 691 (0.1%, [0.1-0.1]) | 285 (0.1%, [0.1-0.1]) | 154 (0.1%, [0.1-0.1]) | 185 (0.1%, [0.1-0.1]) | 1,315 |
| Vaccination status | NA | - | - | 912 (1.2%, [1.1-1.2]) | 2,882 (1.2%, [1.1-1.2]) | 3,794 |
|  | Not vaccinated | - | - | 3,650 (4.6%, [4.5-4.8]) | 9,030 (3.7%, [3.7-3.8]) | 12,680 |
|  | One does | - | - | 519 (0.7%, [0.6-0.7]) | 1,496 (0.6%, [0.6-0.6]) | 2,015 |
|  | Three does | - | - | 14,388 (18.2%, [17.9-18.5]) | 198,035 (81.7%, [81.5-81.8]) | 212,423 |
|  | Two does | - | - | 56,135 (71%, [70.7-71.3]) | 20,246 (8.4%, [8.2-8.5]) | 76,381 |
|  | Unknown does | - | - | 3,433 (4.3%, [4.2-4.5]) | 10,725 (4.4%, [4.3-4.5]) | 14,158 |
| Symptoms | Symptomatic | 95,973 (13.5%, [13.4-13.5]) | 49,228 (13.8%, [13.6-13.9]) | 36,687 (16%, [15.9-16.2]) | 52,983 (21.9%, [21.7-22]) | 234,871 |
| Number of reported symptoms | Mean (SD) | 0.52 (1.66) | 0.5 (1.63) | 0.61 (1.84) | 0.91 (2.34) | 0.6 (1.83) |
| Sought medical attention for symptoms | No | 87,252 (89.5%, [89.3-89.7]) | 43,636 (88.9%, [88.6-89.2]) | 31,933 (87.2%, [86.8-87.5]) | 47,231 (89.2%, [89-89.5]) | 210,052 |
|  | Yes | 10,203 (10.5%, [10.3-10.7]) | 5,457 (11.1%, [10.8-11.4]) | 4,701 (12.8%, [12.5-13.2]) | 5,698 (10.8%, [10.5-11]) | 26,059 |
| Symptoms affecting day-to-day activities | A lot | - | - | 2,291 (1%, [1-1]) | 4,931 (2%, [2-2.1]) | 7,222 |
|  | A little | - | - | 7,849 (3.4%, [3.4-3.5]) | 15,314 (6.3%, [6.2-6.4]) | 23,163 |
|  | Not at all | - | - | 4,938 (2.2%, [2.1-2.2]) | 9,270 (3.8%, [3.7-3.9]) | 14,208 |
|  | Don't know / PNA / Non-reponse | - | - | 21,609 (9.4%, [9.3-9.6]) | 23,468 (9.7%, [9.6-9.8]) | 190,278 |
|  | No symptoms reported | - | - | 192,247 (84%, [83.8-84.1]) | 189,431 (78.1%, [78-78.3]) | 1,307,639 |
| Days since onset of first symptom | Mean (SD) | 7.93 (3.28) | 8.09 (3.28) | 7.89 (3.32) | 7.97 (3.29) | 7.95 (3.29) |
| Symptom in past 7 days | Loss or change of sense of smell | 2,453 (0.3%, [0.3-0.4]) | 1,392 (0.4%, [0.4-0.4]) | 1,460 (0.6%, [0.6-0.7]) | 2,273 (0.9%, [0.9-1]) | 7,578 |
|  | Loss or change of sense of taste | 2,876 (0.4%, [0.4-0.4]) | 1,551 (0.4%, [0.4-0.5]) | 1,420 (0.6%, [0.6-0.7]) | 2,362 (1%, [0.9-1]) | 8,209 |
|  | New persistent cough | 4,710 (0.7%, [0.6-0.7]) | 2,120 (0.6%, [0.6-0.6]) | 3,599 (1.6%, [1.5-1.6]) | 7,212 (3%, [2.9-3]) | 17,641 |
|  | Fever | 5,666 (0.8%, [0.8-0.8]) | 2,515 (0.7%, [0.7-0.7]) | 2,619 (1.1%, [1.1-1.2]) | 4,450 (1.8%, [1.8-1.9]) | 15,250 |
|  | Runny nose | 24,262 (3.4%, [3.4-3.4]) | 12,008 (3.4%, [3.3-3.4]) | 11,105 (4.9%, [4.8-4.9]) | 19,335 (8%, [7.9-8.1]) | 66,710 |
|  | Sneezing | 24,496 (3.4%, [3.4-3.5]) | 10,909 (3%, [3-3.1]) | 9,673 (4.2%, [4.1-4.3]) | 15,087 (6.2%, [6.1-6.3]) | 60,165 |
|  | Blocked nose | 17,806 (2.5%, [2.5-2.5]) | 8,504 (2.4%, [2.3-2.4]) | 8,067 (3.5%, [3.4-3.6]) | 13,507 (5.6%, [5.5-5.7]) | 47,884 |
|  | Sore eyes | 14,341 (2%, [2-2]) | 6,777 (1.9%, [1.8-1.9]) | 4,628 (2%, [2-2.1]) | 7,038 (2.9%, [2.8-3]) | 32,784 |

|  |  |  |  |  |  |
| --- | --- | --- | --- | --- | --- |
| Sore throat | 27,295 (3.8%, [3.8-3.9]) | 10,259 (2.9%, [2.8-2.9]) | 10,410 (4.5%, [4.5-4.6]) | 15,571 (6.4%, [6.3-6.5]) | 63,535 |
| Hoarse voice | 8,322 (1.2%, [1.1-1.2]) | 3,292 (0.9%, [0.9-1]) | 4,509 (2%, [1.9-2]) | 7,247 (3%, [2.9-3.1]) | 23,370 |
| Headache | 39,332 (5.5%, [5.5-5.6]) | 20,835 (5.8%, [5.7-5.9]) | 13,727 (6%, [5.9-6.1]) | 21,475 (8.9%, [8.7-9]) | 95,369 |
| Dizziness | 12,857 (1.8%, [1.8-1.8]) | 6,128 (1.7%, [1.7-1.8]) | 4,372 (1.9%, [1.9-2]) | 6,307 (2.6%, [2.5-2.7]) | 29,664 |
| Appetite loss | 6,676 (0.9%, [0.9-1]) | 3,318 (0.9%, [0.9-1]) | 2,736 (1.2%, [1.2-1.2]) | 4,296 (1.8%, [1.7-1.8]) | 17,026 |
| Nausea/vomiting | 8,185 (1.1%, [1.1-1.2]) | 3,884 (1.1%, [1.1-1.1]) | 2,825 (1.2%, [1.2-1.3]) | 3,948 (1.6%, [1.6-1.7]) | 18,842 |
| Diarrhoea | 11,107 (1.6%, [1.5-1.6]) | 4,799 (1.3%, [1.3-1.4]) | 3,573 (1.6%, [1.5-1.6]) | 4,741 (2%, [1.9-2]) | 24,220 |
| Abdominal pain / belly ache | 14,707 (2.1%, [2-2.1]) | 6,934 (1.9%, [1.9-2]) | 4,619 (2%, [2-2.1]) | 6,096 (2.5%, [2.5-2.6]) | 32,356 |
| Shortness of breath | 12,907 (1.8%, [1.8-1.8]) | 6,634 (1.9%, [1.8-1.9]) | 4,587 (2%, [1.9-2.1]) | 7,647 (3.2%, [3.1-3.2]) | 31,775 |
| Tight chest | 8,977 (1.3%, [1.2-1.3]) | 4,514 (1.3%, [1.2-1.3]) | 2,969 (1.3%, [1.3-1.3]) | 4,736 (2%, [1.9-2]) | 21,196 |
| Chest pain | 4,158 (0.6%, [0.6-0.6]) | 2,279 (0.6%, [0.6-0.7]) | 1,529 (0.7%, [0.6-0.7]) | 2,494 (1%, [1-1.1]) | 10,460 |
| Chills | 7,077 (1%, [1-1]) | 5,429 (1.5%, [1.5-1.6]) | 2,652 (1.2%, [1.1-1.2]) | 5,955 (2.5%, [2.4-2.5]) | 21,113 |
| Difficulty sleeping | 26,551 (3.7%, [3.7-3.8]) | 14,043 (3.9%, [3.9-4]) | 8,956 (3.9%, [3.8-4]) | 13,035 (5.4%, [5.3-5.5]) | 62,585 |
| Tiredness | 39,897 (5.6%, [5.5-5.6]) | 19,756 (5.5%, [5.4-5.6]) | 14,813 (6.5%, [6.4-6.6]) | 23,072 (9.5%, [9.4-9.6]) | 97,538 |
| Severe fatigue | 4,677 (0.7%, [0.6-0.7]) | 2,715 (0.8%, [0.7-0.8]) | 2,063 (0.9%, [0.9-0.9]) | 3,482 (1.4%, [1.4-1.5]) | 12,937 |
| Numbness/tingling | 9,748 (1.4%, [1.3-1.4]) | 4,725 (1.3%, [1.3-1.4]) | 3,214 (1.4%, [1.4-1.5]) | 4,295 (1.8%, [1.7-1.8]) | 21,982 |
| Heavy arms/legs | 8,527 (1.2%, [1.2-1.2]) | 4,017 (1.1%, [1.1-1.2]) | 2,707 (1.2%, [1.1-1.2]) | 4,080 (1.7%, [1.6-1.7]) | 19,331 |
| Muscle aches | 21,342 (3%, [3-3]) | 11,069 (3.1%, [3-3.1]) | 7,488 (3.3%, [3.2-3.3]) | 11,866 (4.9%, [4.8-5]) | 51,765 |

**Table S2.**
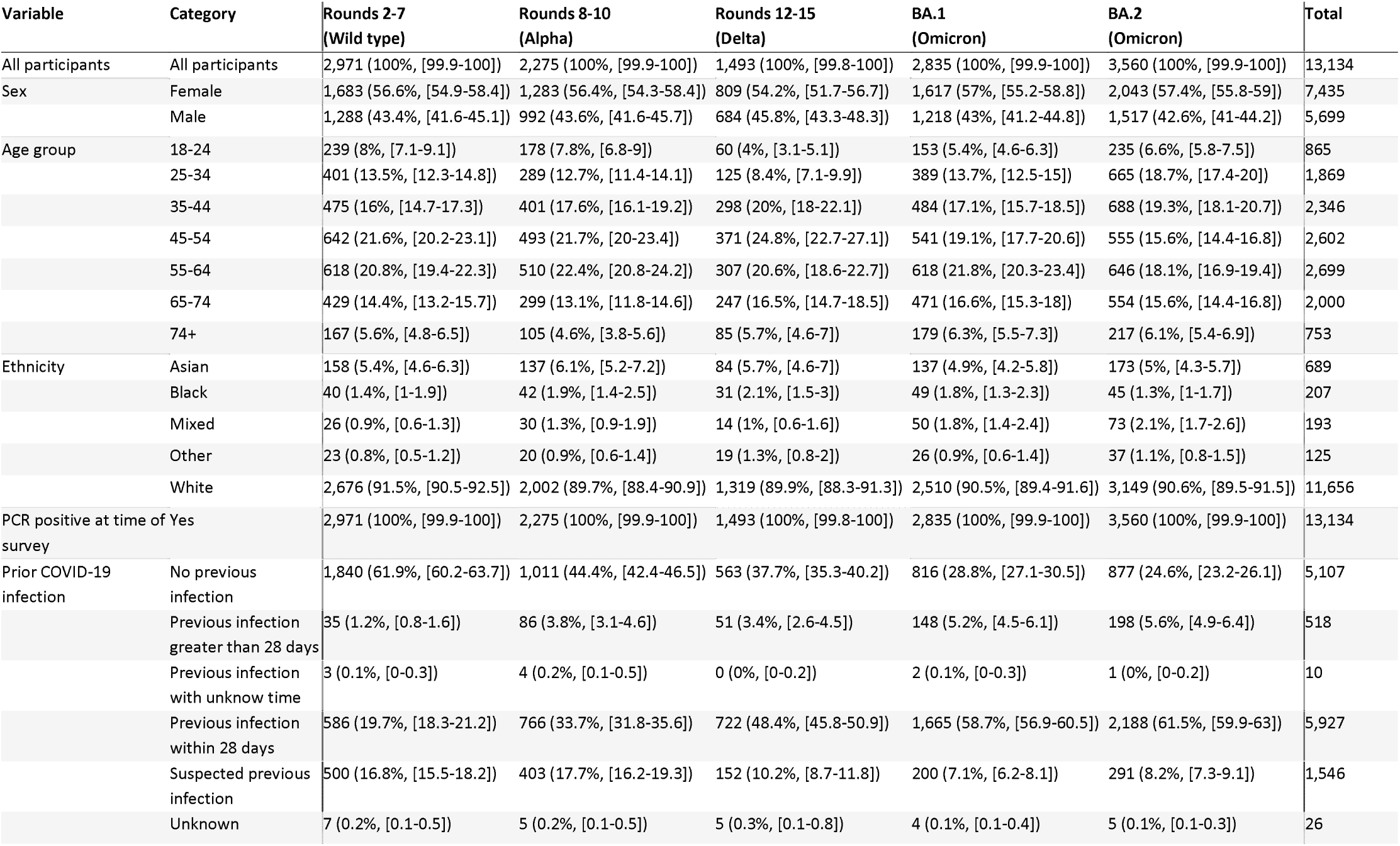
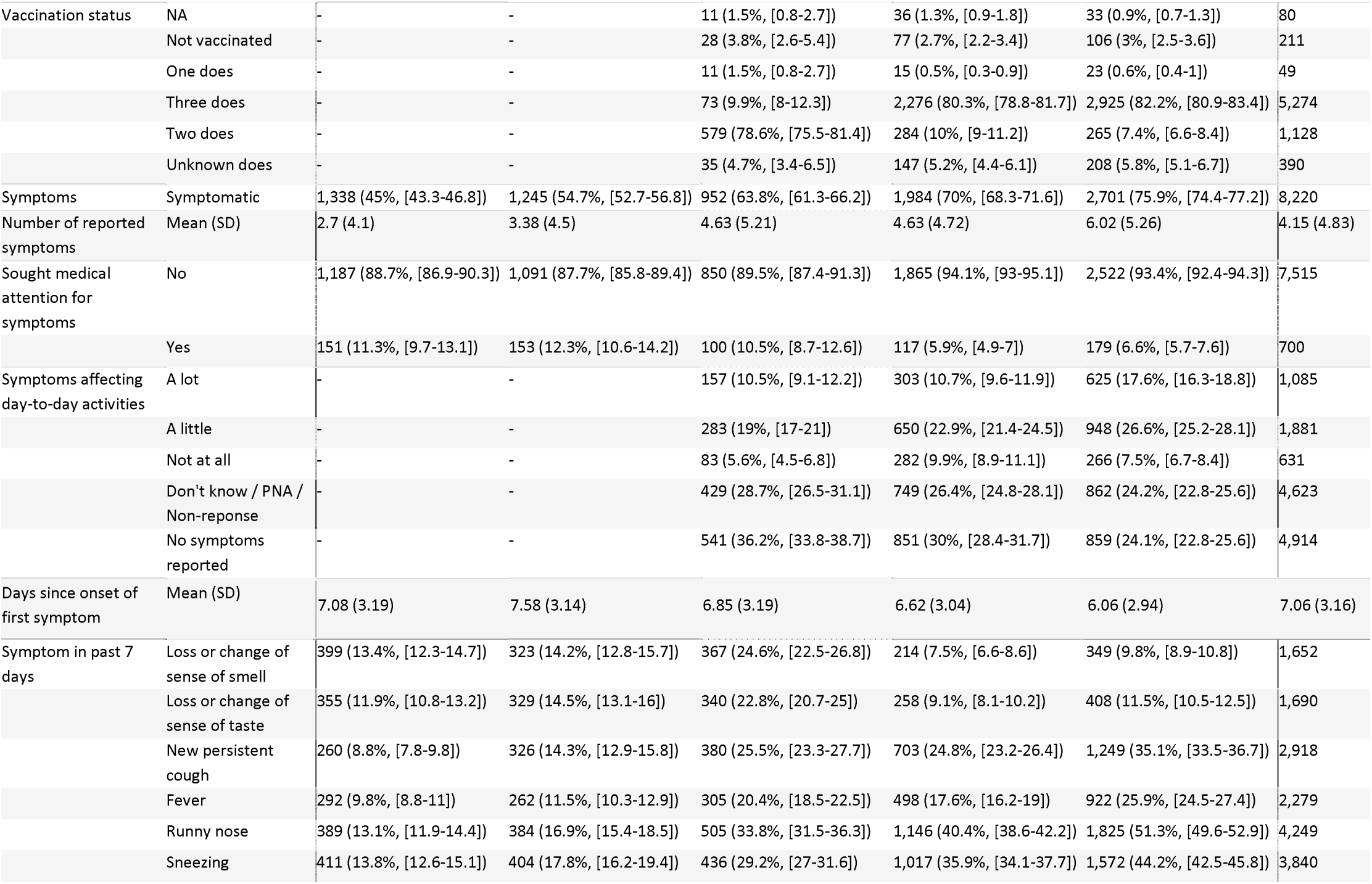
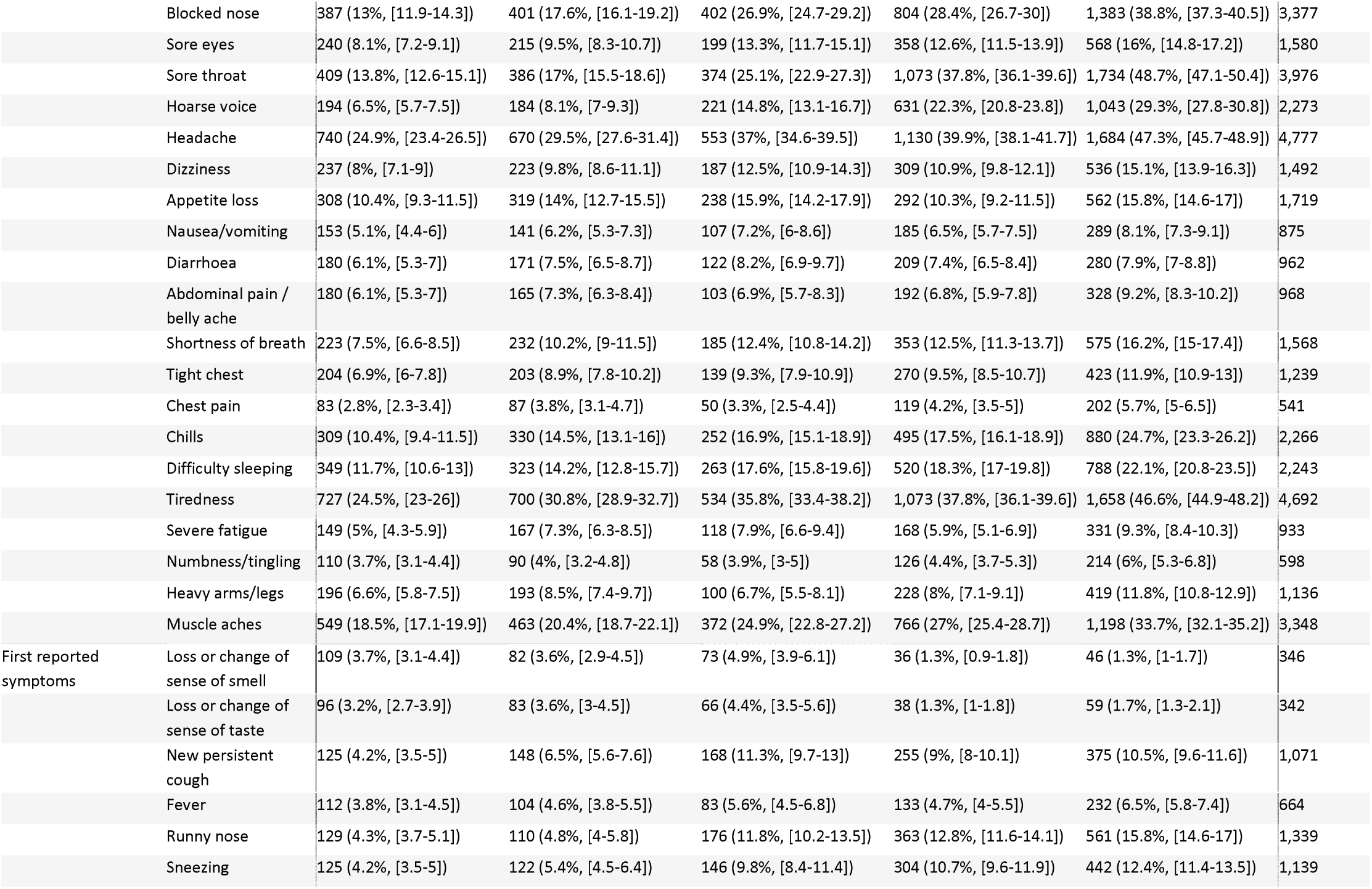
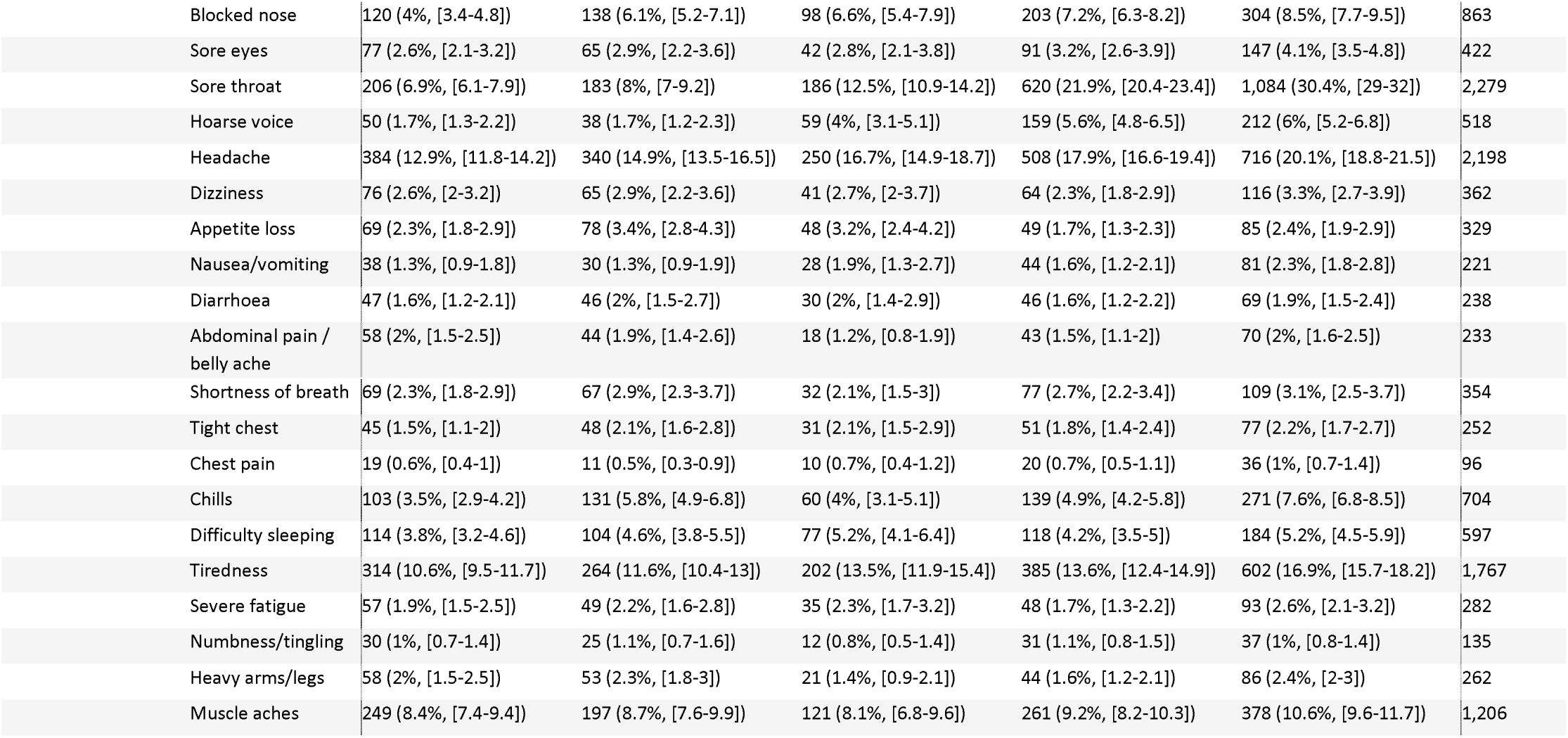
Characteristics of swab positives in study population

| Variable | Category | Rounds 2-7<br>(Wild type) | Rounds 8-10<br>(Alpha) | Rounds 12-15<br>(Delta) | BA.1<br>(Omicron) | BA.2<br>(Omicron) | Total |
| --- | --- | --- | --- | --- | --- | --- | --- |
| All participants | All participants | 2,971 (100%, [99.9-100]) | 2,275 (100%, [99.9-100]) | 1,493 (100%, [99.8-100]) | 2,835 (100%, [99.9-100]) | 3,560 (100%, [99.9-100]) | 13,134 |
| Sex | Female | 1,683 (56.6%, [54.9-58.4]) | 1,283 (56.4%, [54.3-58.4]) | 809 (54.2%, [51.7-56.7]) | 1,617 (57%, [55.2-58.8]) | 2,043 (57.4%, [55.8-59]) | 7,435 |
|  | Male | 1,288 (43.4%, [41.6-45.1]) | 992 (43.6%, [41.6-45.7]) | 684 (45.8%, [43.3-48.3]) | 1,218 (43%, [41.2-44.8]) | 1,517 (42.6%, [41-44.2]) | 5,699 |
| Age group | 18-24 | 239 (8%, [7.1-9.1]) | 178 (7.8%, [6.8-9]) | 60 (4%, [3.1-5.1]) | 153 (5.4%, [4.6-6.3]) | 235 (6.6%, [5.8-7.5]) | 865 |
|  | 25-34 | 401 (13.5%, [12.3-14.8]) | 289 (12.7%, [11.4-14.1]) | 125 (8.4%, [7.1-9.9]) | 389 (13.7%, [12.5-15]) | 665 (18.7%, [17.4-20]) | 1,869 |
|  | 35-44 | 475 (16%, [14.7-17.3]) | 401 (17.6%, [16.1-19.2]) | 298 (20%, [18-22.1]) | 484 (17.1%, [15.7-18.5]) | 688 (19.3%, [18.1-20.7]) | 2,346 |
|  | 45-54 | 642 (21.6%, [20.2-23.1]) | 493 (21.7%, [20-23.4]) | 371 (24.8%, [22.7-27.1]) | 541 (19.1%, [17.7-20.6]) | 555 (15.6%, [14.4-16.8]) | 2,602 |
|  | 55-64 | 618 (20.8%, [19.4-22.3]) | 510 (22.4%, [20.8-24.2]) | 307 (20.6%, [18.6-22.7]) | 618 (21.8%, [20.3-23.4]) | 646 (18.1%, [16.9-19.4]) | 2,699 |
|  | 65-74 | 429 (14.4%, [13.2-15.7]) | 299 (13.1%, [11.8-14.6]) | 247 (16.5%, [14.7-18.5]) | 471 (16.6%, [15.3-18]) | 554 (15.6%, [14.4-16.8]) | 2,000 |
|  | 74+ | 167 (5.6%, [4.8-6.5]) | 105 (4.6%, [3.8-5.6]) | 85 (5.7%, [4.6-7]) | 179 (6.3%, [5.5-7.3]) | 217 (6.1%, [5.4-6.9]) | 753 |
| Ethnicity | Asian | 158 (5.4%, [4.6-6.3]) | 137 (6.1%, [5.2-7.2]) | 84 (5.7%, [4.6-7]) | 137 (4.9%, [4.2-5.8]) | 173 (5%, [4.3-5.7]) | 689 |
|  | Black | 40 (1.4%, [1-1.9]) | 42 (1.9%, [1.4-2.5]) | 31 (2.1%, [1.5-3]) | 49 (1.8%, [1.3-2.3]) | 45 (1.3%, [1-1.7]) | 207 |
|  | Mixed | 26 (0.9%, [0.6-1.3]) | 30 (1.3%, [0.9-1.9]) | 14 (1%, [0.6-1.6]) | 50 (1.8%, [1.4-2.4]) | 73 (2.1%, [1.7-2.6]) | 193 |
|  | Other | 23 (0.8%, [0.5-1.2]) | 20 (0.9%, [0.6-1.4]) | 19 (1.3%, [0.8-2]) | 26 (0.9%, [0.6-1.4]) | 37 (1.1%, [0.8-1.5]) | 125 |
|  | White | 2,676 (91.5%, [90.5-92.5]) | 2,002 (89.7%, [88.4-90.9]) | 1,319 (89.9%, [88.3-91.3]) | 2,510 (90.5%, [89.4-91.6]) | 3,149 (90.6%, [89.5-91.5]) | 11,656 |
| PCR positive at time of survey | Yes | 2,971 (100%, [99.9-100]) | 2,275 (100%, [99.9-100]) | 1,493 (100%, [99.8-100]) | 2,835 (100%, [99.9-100]) | 3,560 (100%, [99.9-100]) | 13,134 |
| Prior COVID-19 infection | No previous infection | 1,840 (61.9%, [60.2-63.7]) | 1,011 (44.4%, [42.4-46.5]) | 563 (37.7%, [35.3-40.2]) | 816 (28.8%, [27.1-30.5]) | 877 (24.6%, [23.2-26.1]) | 5,107 |
|  | Previous infection greater than 28 days | 35 (1.2%, [0.8-1.6]) | 86 (3.8%, [3.1-4.6]) | 51 (3.4%, [2.6-4.5]) | 148 (5.2%, [4.5-6.1]) | 198 (5.6%, [4.9-6.4]) | 518 |
|  | Previous infection with unknow time | 3 (0.1%, [0-0.3]) | 4 (0.2%, [0.1-0.5]) | 0 (0%, [0-0.2]) | 2 (0.1%, [0-0.3]) | 1 (0%, [0-0.2]) | 10 |
|  | Previous infection within 28 days | 586 (19.7%, [18.3-21.2]) | 766 (33.7%, [31.8-35.6]) | 722 (48.4%, [45.8-50.9]) | 1,665 (58.7%, [56.9-60.5]) | 2,188 (61.5%, [59.9-63]) | 5,927 |
|  | Suspected previous infection | 500 (16.8%, [15.5-18.2]) | 403 (17.7%, [16.2-19.3]) | 152 (10.2%, [8.7-11.8]) | 200 (7.1%, [6.2-8.1]) | 291 (8.2%, [7.3-9.1]) | 1,546 |
|  | Unknown | 7 (0.2%, [0.1-0.5]) | 5 (0.2%, [0.1-0.5]) | 5 (0.3%, [0.1-0.8]) | 4 (0.1%, [0.1-0.4]) | 5 (0.1%, [0.1-0.3]) | 26 |
| Vaccination status | NA | - | - | 11 (1.5%, [0.8-2.7]) | 36 (1.3%, [0.9-1.8]) | 33 (0.9%, [0.7-1.3]) | 80 |
|  | Not vaccinated | - | - | 28 (3.8%, [2.6-5.4]) | 77 (2.7%, [2.2-3.4]) | 106 (3%, [2.5-3.6]) | 211 |
|  | One does | - | - | 11 (1.5%, [0.8-2.7]) | 15 (0.5%, [0.3-0.9]) | 23 (0.6%, [0.4-1]) | 49 |
|  | Three does | - | - | 73 (9.9%, [8-12.3]) | 2,276 (80.3%, [78.8-81.7]) | 2,925 (82.2%, [80.9-83.4]) | 5,274 |
|  | Two does | - | - | 579 (78.6%, [75.5-81.4]) | 284 (10%, [9-11.2]) | 265 (7.4%, [6.6-8.4]) | 1,128 |
|  | Unknown does | - | - | 35 (4.7%, [3.4-6.5]) | 147 (5.2%, [4.4-6.1]) | 208 (5.8%, [5.1-6.7]) | 390 |
| Symptoms | Symptomatic | 1,338 (45%, [43.3-46.8]) | 1,245 (54.7%, [52.7-56.8]) | 952 (63.8%, [61.3-66.2]) | 1,984 (70%, [68.3-71.6]) | 2,701 (75.9%, [74.4-77.2]) | 8,220 |
| Number of reported symptoms | Mean (SD) | 2.7 (4.1) | 3.38 (4.5) | 4.63 (5.21) | 4.63 (4.72) | 6.02 (5.26) | 4.15 (4.83) |
| Sought medical attention for symptoms | No | 1,187 (88.7%, [86.9-90.3]) | 1,091 (87.7%, [85.8-89.4]) | 850 (89.5%, [87.4-91.3]) | 1,865 (94.1%, [93-95.1]) | 2,522 (93.4%, [92.4-94.3]) | 7,515 |
|  | Yes | 151 (11.3%, [9.7-13.1]) | 153 (12.3%, [10.6-14.2]) | 100 (10.5%, [8.7-12.6]) | 117 (5.9%, [4.9-7]) | 179 (6.6%, [5.7-7.6]) | 700 |
| Symptoms affecting day-to-day activities | A lot | - | - | 157 (10.5%, [9.1-12.2]) | 303 (10.7%, [9.6-11.9]) | 625 (17.6%, [16.3-18.8]) | 1,085 |
|  | A little | - | - | 283 (19%, [17-21]) | 650 (22.9%, [21.4-24.5]) | 948 (26.6%, [25.2-28.1]) | 1,881 |
|  | Not at all | - | - | 83 (5.6%, [4.5-6.8]) | 282 (9.9%, [8.9-11.1]) | 266 (7.5%, [6.7-8.4]) | 631 |
|  | Don't know / PNA / Non-reponse | - | - | 429 (28.7%, [26.5-31.1]) | 749 (26.4%, [24.8-28.1]) | 862 (24.2%, [22.8-25.6]) | 4,623 |
|  | No symptoms reported | - | - | 541 (36.2%, [33.8-38.7]) | 851 (30%, [28.4-31.7]) | 859 (24.1%, [22.8-25.6]) | 4,914 |
| Days since onset of first symptom | Mean (SD) | 7.08 (3.19) | 7.58 (3.14) | 6.85 (3.19) | 6.62 (3.04) | 6.06 (2.94) | 7.06 (3.16) |
| Symptom in past 7 days | Loss or change of sense of smell | 399 (13.4%, [12.3-14.7]) | 323 (14.2%, [12.8-15.7]) | 367 (24.6%, [22.5-26.8]) | 214 (7.5%, [6.6-8.6]) | 349 (9.8%, [8.9-10.8]) | 1,652 |
|  | Loss or change of sense of taste | 355 (11.9%, [10.8-13.2]) | 329 (14.5%, [13.1-16]) | 340 (22.8%, [20.7-25]) | 258 (9.1%, [8.1-10.2]) | 408 (11.5%, [10.5-12.5]) | 1,690 |
|  | New persistent cough | 260 (8.8%, [7.8-9.8]) | 326 (14.3%, [12.9-15.8]) | 380 (25.5%, [23.3-27.7]) | 703 (24.8%, [23.2-26.4]) | 1,249 (35.1%, [33.5-36.7]) | 2,918 |
|  | Fever | 292 (9.8%, [8.8-11]) | 262 (11.5%, [10.3-12.9]) | 305 (20.4%, [18.5-22.5]) | 498 (17.6%, [16.2-19]) | 922 (25.9%, [24.5-27.4]) | 2,279 |
|  | Runny nose | 389 (13.1%, [11.9-14.4]) | 384 (16.9%, [15.4-18.5]) | 505 (33.8%, [31.5-36.3]) | 1,146 (40.4%, [38.6-42.2]) | 1,825 (51.3%, [49.6-52.9]) | 4,249 |
|  | Sneezing | 411 (13.8%, [12.6-15.1]) | 404 (17.8%, [16.2-19.4]) | 436 (29.2%, [27-31.6]) | 1,017 (35.9%, [34.1-37.7]) | 1,572 (44.2%, [42.5-45.8]) | 3,840 |
|  | Blocked nose | 387 (13%, [11.9-14.3]) | 401 (17.6%, [16.1-19.2]) | 402 (26.9%, [24.7-29.2]) | 804 (28.4%, [26.7-30]) | 1,383 (38.8%, [37.3-40.5]) | 3,377 |
|  | Sore eyes | 240 (8.1%, [7.2-9.1]) | 215 (9.5%, [8.3-10.7]) | 199 (13.3%, [11.7-15.1]) | 358 (12.6%, [11.5-13.9]) | 568 (16%, [14.8-17.2]) | 1,580 |
|  | Sore throat | 409 (13.8%, [12.6-15.1]) | 386 (17%, [15.5-18.6]) | 374 (25.1%, [22.9-27.3]) | 1,073 (37.8%, [36.1-39.6]) | 1,734 (48.7%, [47.1-50.4]) | 3,976 |
|  | Hoarse voice | 194 (6.5%, [5.7-7.5]) | 184 (8.1%, [7-9.3]) | 221 (14.8%, [13.1-16.7]) | 631 (22.3%, [20.8-23.8]) | 1,043 (29.3%, [27.8-30.8]) | 2,273 |
|  | Headache | 740 (24.9%, [23.4-26.5]) | 670 (29.5%, [27.6-31.4]) | 553 (37%, [34.6-39.5]) | 1,130 (39.9%, [38.1-41.7]) | 1,684 (47.3%, [45.7-48.9]) | 4,777 |
|  | Dizziness | 237 (8%, [7.1-9]) | 223 (9.8%, [8.6-11.1]) | 187 (12.5%, [10.9-14.3]) | 309 (10.9%, [9.8-12.1]) | 536 (15.1%, [13.9-16.3]) | 1,492 |
|  | Appetite loss | 308 (10.4%, [9.3-11.5]) | 319 (14%, [12.7-15.5]) | 238 (15.9%, [14.2-17.9]) | 292 (10.3%, [9.2-11.5]) | 562 (15.8%, [14.6-17]) | 1,719 |
|  | Nausea/vomiting | 153 (5.1%, [4.4-6]) | 141 (6.2%, [5.3-7.3]) | 107 (7.2%, [6-8.6]) | 185 (6.5%, [5.7-7.5]) | 289 (8.1%, [7.3-9.1]) | 875 |
|  | Diarrhoea | 180 (6.1%, [5.3-7]) | 171 (7.5%, [6.5-8.7]) | 122 (8.2%, [6.9-9.7]) | 209 (7.4%, [6.5-8.4]) | 280 (7.9%, [7-8.8]) | 962 |
|  | Abdominal pain /<br>belly ache | 180 (6.1%, [5.3-7]) | 165 (7.3%, [6.3-8.4]) | 103 (6.9%, [5.7-8.3]) | 192 (6.8%, [5.9-7.8]) | 328 (9.2%, [8.3-10.2]) | 968 |
|  | Shortness of breath | 223 (7.5%, [6.6-8.5]) | 232 (10.2%, [9-11.5]) | 185 (12.4%, [10.8-14.2]) | 353 (12.5%, [11.3-13.7]) | 575 (16.2%, [15-17.4]) | 1,568 |
|  | Tight chest | 204 (6.9%, [6-7.8]) | 203 (8.9%, [7.8-10.2]) | 139 (9.3%, [7.9-10.9]) | 270 (9.5%, [8.5-10.7]) | 423 (11.9%, [10.9-13]) | 1,239 |
|  | Chest pain | 83 (2.8%, [2.3-3.4]) | 87 (3.8%, [3.1-4.7]) | 50 (3.3%, [2.5-4.4]) | 119 (4.2%, [3.5-5]) | 202 (5.7%, [5-6.5]) | 541 |
|  | Chills | 309 (10.4%, [9.4-11.5]) | 330 (14.5%, [13.1-16]) | 252 (16.9%, [15.1-18.9]) | 495 (17.5%, [16.1-18.9]) | 880 (24.7%, [23.3-26.2]) | 2,266 |
|  | Difficulty sleeping | 349 (11.7%, [10.6-13]) | 323 (14.2%, [12.8-15.7]) | 263 (17.6%, [15.8-19.6]) | 520 (18.3%, [17-19.8]) | 788 (22.1%, [20.8-23.5]) | 2,243 |
|  | Tiredness | 727 (24.5%, [23-26]) | 700 (30.8%, [28.9-32.7]) | 534 (35.8%, [33.4-38.2]) | 1,073 (37.8%, [36.1-39.6]) | 1,658 (46.6%, [44.9-48.2]) | 4,692 |
|  | Severe fatigue | 149 (5%, [4.3-5.9]) | 167 (7.3%, [6.3-8.5]) | 118 (7.9%, [6.6-9.4]) | 168 (5.9%, [5.1-6.9]) | 331 (9.3%, [8.4-10.3]) | 933 |
|  | Numbness/tingling | 110 (3.7%, [3.1-4.4]) | 90 (4%, [3.2-4.8]) | 58 (3.9%, [3-5]) | 126 (4.4%, [3.7-5.3]) | 214 (6%, [5.3-6.8]) | 598 |
|  | Heavy arms/legs | 196 (6.6%, [5.8-7.5]) | 193 (8.5%, [7.4-9.7]) | 100 (6.7%, [5.5-8.1]) | 228 (8%, [7.1-9.1]) | 419 (11.8%, [10.8-12.9]) | 1,136 |
|  | Muscle aches | 549 (18.5%, [17.1-19.9]) | 463 (20.4%, [18.7-22.1]) | 372 (24.9%, [22.8-27.2]) | 766 (27%, [25.4-28.7]) | 1,198 (33.7%, [32.1-35.2]) | 3,348 |
| First reported<br>symptoms | Loss or change of<br>sense of smell | 109 (3.7%, [3.1-4.4]) | 82 (3.6%, [2.9-4.5]) | 73 (4.9%, [3.9-6.1]) | 36 (1.3%, [0.9-1.8]) | 46 (1.3%, [1-1.7]) | 346 |
|  | Loss or change of<br>sense of taste | 96 (3.2%, [2.7-3.9]) | 83 (3.6%, [3-4.5]) | 66 (4.4%, [3.5-5.6]) | 38 (1.3%, [1-1.8]) | 59 (1.7%, [1.3-2.1]) | 342 |
|  | New persistent<br>cough | 125 (4.2%, [3.5-5]) | 148 (6.5%, [5.6-7.6]) | 168 (11.3%, [9.7-13]) | 255 (9%, [8-10.1]) | 375 (10.5%, [9.6-11.6]) | 1,071 |
|  | Fever | 112 (3.8%, [3.1-4.5]) | 104 (4.6%, [3.8-5.5]) | 83 (5.6%, [4.5-6.8]) | 133 (4.7%, [4-5.5]) | 232 (6.5%, [5.8-7.4]) | 664 |
|  | Runny nose | 129 (4.3%, [3.7-5.1]) | 110 (4.8%, [4-5.8]) | 176 (11.8%, [10.2-13.5]) | 363 (12.8%, [11.6-14.1]) | 561 (15.8%, [14.6-17]) | 1,339 |
|  | Sneezing | 125 (4.2%, [3.5-5]) | 122 (5.4%, [4.5-6.4]) | 146 (9.8%, [8.4-11.4]) | 304 (10.7%, [9.6-11.9]) | 442 (12.4%, [11.4-13.5]) | 1,139 |

|  |  |  |  |  |  |  |
| --- | --- | --- | --- | --- | --- | --- |
| Blocked nose | 120 (4%, [3.4-4.8]) | 138 (6.1%, [5.2-7.1]) | 98 (6.6%, [5.4-7.9]) | 203 (7.2%, [6.3-8.2]) | 304 (8.5%, [7.7-9.5]) | 863 |
| Sore eyes | 77 (2.6%, [2.1-3.2]) | 65 (2.9%, [2.2-3.6]) | 42 (2.8%, [2.1-3.8]) | 91 (3.2%, [2.6-3.9]) | 147 (4.1%, [3.5-4.8]) | 422 |
| Sore throat | 206 (6.9%, [6.1-7.9]) | 183 (8%, [7-9.2]) | 186 (12.5%, [10.9-14.2]) | 620 (21.9%, [20.4-23.4]) | 1,084 (30.4%, [29-32]) | 2,279 |
| Hoarse voice | 50 (1.7%, [1.3-2.2]) | 38 (1.7%, [1.2-2.3]) | 59 (4%, [3.1-5.1]) | 159 (5.6%, [4.8-6.5]) | 212 (6%, [5.2-6.8]) | 518 |
| Headache | 384 (12.9%, [11.8-14.2]) | 340 (14.9%, [13.5-16.5]) | 250 (16.7%, [14.9-18.7]) | 508 (17.9%, [16.6-19.4]) | 716 (20.1%, [18.8-21.5]) | 2,198 |
| Dizziness | 76 (2.6%, [2-3.2]) | 65 (2.9%, [2.2-3.6]) | 41 (2.7%, [2-3.7]) | 64 (2.3%, [1.8-2.9]) | 116 (3.3%, [2.7-3.9]) | 362 |
| Appetite loss | 69 (2.3%, [1.8-2.9]) | 78 (3.4%, [2.8-4.3]) | 48 (3.2%, [2.4-4.2]) | 49 (1.7%, [1.3-2.3]) | 85 (2.4%, [1.9-2.9]) | 329 |
| Nausea/vomiting | 38 (1.3%, [0.9-1.8]) | 30 (1.3%, [0.9-1.9]) | 28 (1.9%, [1.3-2.7]) | 44 (1.6%, [1.2-2.1]) | 81 (2.3%, [1.8-2.8]) | 221 |
| Diarrhoea | 47 (1.6%, [1.2-2.1]) | 46 (2%, [1.5-2.7]) | 30 (2%, [1.4-2.9]) | 46 (1.6%, [1.2-2.2]) | 69 (1.9%, [1.5-2.4]) | 238 |
| Abdominal pain /<br>belly ache | 58 (2%, [1.5-2.5]) | 44 (1.9%, [1.4-2.6]) | 18 (1.2%, [0.8-1.9]) | 43 (1.5%, [1.1-2]) | 70 (2%, [1.6-2.5]) | 233 |
| Shortness of breath | 69 (2.3%, [1.8-2.9]) | 67 (2.9%, [2.3-3.7]) | 32 (2.1%, [1.5-3]) | 77 (2.7%, [2.2-3.4]) | 109 (3.1%, [2.5-3.7]) | 354 |
| Tight chest | 45 (1.5%, [1.1-2]) | 48 (2.1%, [1.6-2.8]) | 31 (2.1%, [1.5-2.9]) | 51 (1.8%, [1.4-2.4]) | 77 (2.2%, [1.7-2.7]) | 252 |
| Chest pain | 19 (0.6%, [0.4-1]) | 11 (0.5%, [0.3-0.9]) | 10 (0.7%, [0.4-1.2]) | 20 (0.7%, [0.5-1.1]) | 36 (1%, [0.7-1.4]) | 96 |
| Chills | 103 (3.5%, [2.9-4.2]) | 131 (5.8%, [4.9-6.8]) | 60 (4%, [3.1-5.1]) | 139 (4.9%, [4.2-5.8]) | 271 (7.6%, [6.8-8.5]) | 704 |
| Difficulty sleeping | 114 (3.8%, [3.2-4.6]) | 104 (4.6%, [3.8-5.5]) | 77 (5.2%, [4.1-6.4]) | 118 (4.2%, [3.5-5]) | 184 (5.2%, [4.5-5.9]) | 597 |
| Tiredness | 314 (10.6%, [9.5-11.7]) | 264 (11.6%, [10.4-13]) | 202 (13.5%, [11.9-15.4]) | 385 (13.6%, [12.4-14.9]) | 602 (16.9%, [15.7-18.2]) | 1,767 |
| Severe fatigue | 57 (1.9%, [1.5-2.5]) | 49 (2.2%, [1.6-2.8]) | 35 (2.3%, [1.7-3.2]) | 48 (1.7%, [1.3-2.2]) | 93 (2.6%, [2.1-3.2]) | 282 |
| Numbness/tingling | 30 (1%, [0.7-1.4]) | 25 (1.1%, [0.7-1.6]) | 12 (0.8%, [0.5-1.4]) | 31 (1.1%, [0.8-1.5]) | 37 (1%, [0.8-1.4]) | 135 |
| Heavy arms/legs | 58 (2%, [1.5-2.5]) | 53 (2.3%, [1.8-3]) | 21 (1.4%, [0.9-2.1]) | 44 (1.6%, [1.2-2.1]) | 86 (2.4%, [2-3]) | 262 |
| Muscle aches | 249 (8.4%, [7.4-9.4]) | 197 (8.7%, [7.6-9.9]) | 121 (8.1%, [6.8-9.6]) | 261 (9.2%, [8.2-10.3]) | 378 (10.6%, [9.6-11.7]) | 1,206 |

**Table S3.**
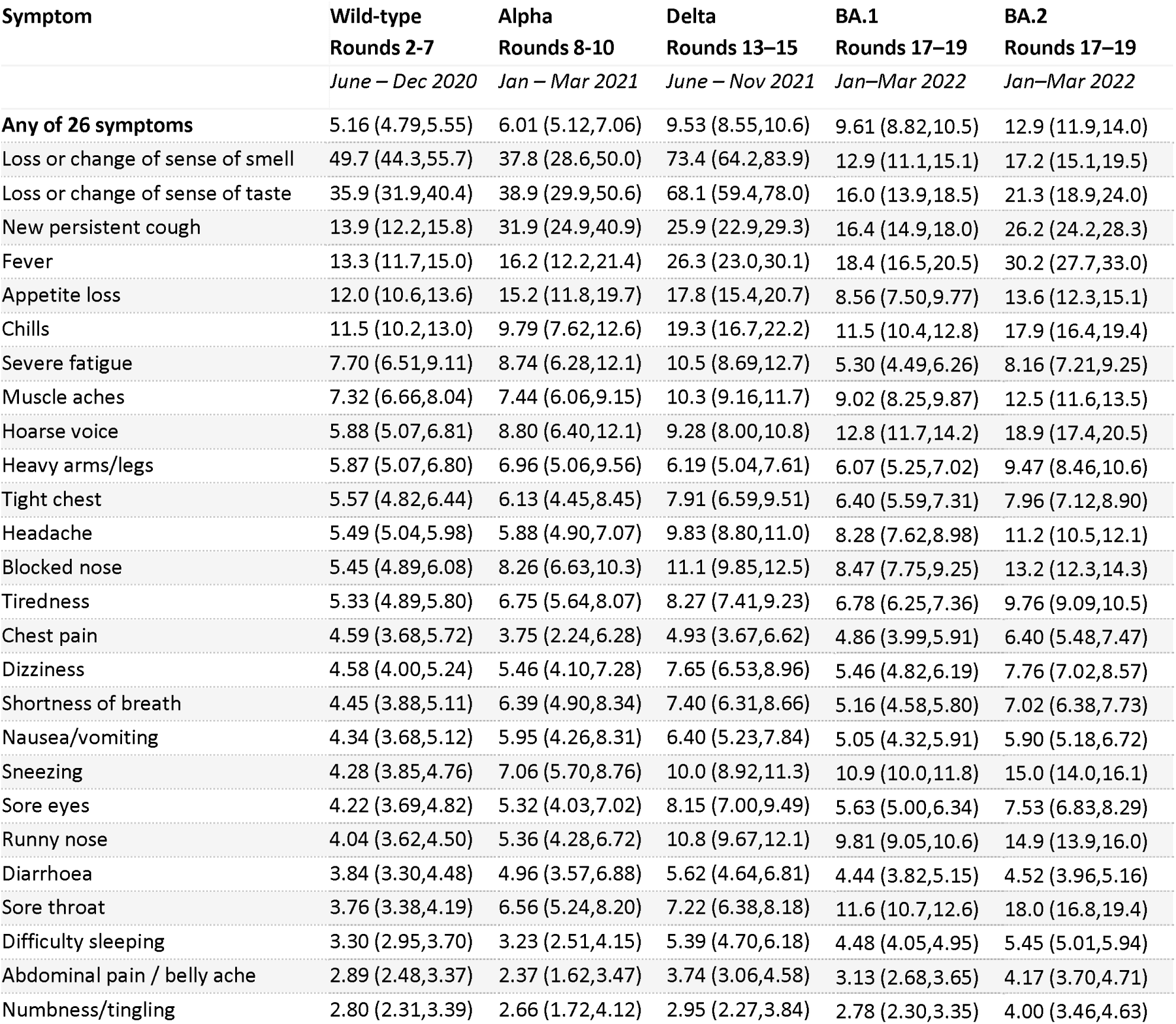
Odds ratios for swab positivity based on presence or absence of any of 26 symptoms surveyed across selected rounds of REACT-1. ORs are derived from logistic regression models with swab positive (1/0) as the outcome variable, adjusted on age, sex and vaccination status.

**Figure S1.**
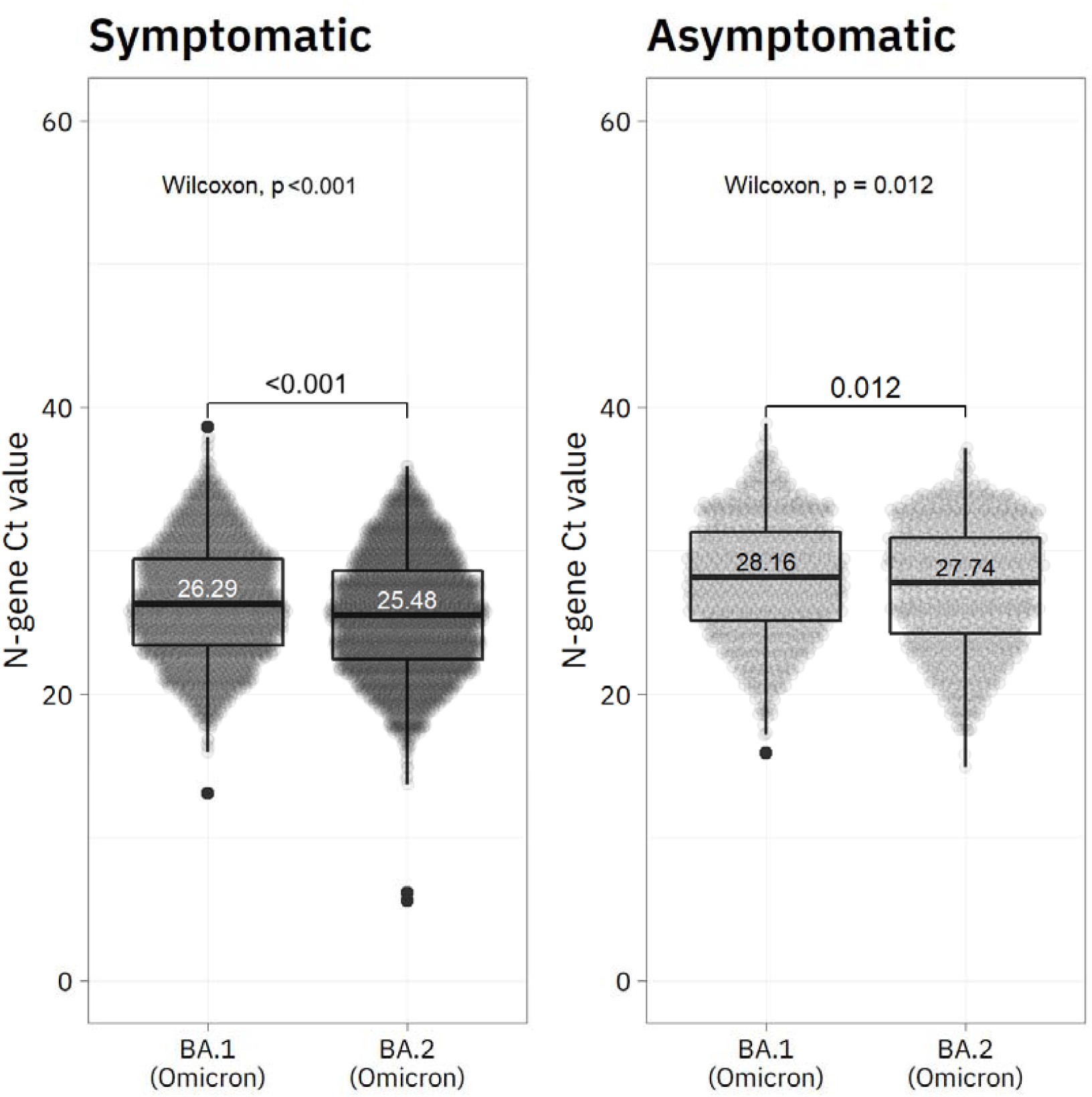
Comparison of Ct values in swab-positive respondents with Omicron BA.1 and BA.2 variants in REACT-1 rounds 17–19, stratified by symptom status (presence of any of 26 surveyed symptoms). Ct values are lower in BA.2, in both asymptomatic and symptomatic respondents.

**Figure S2.**
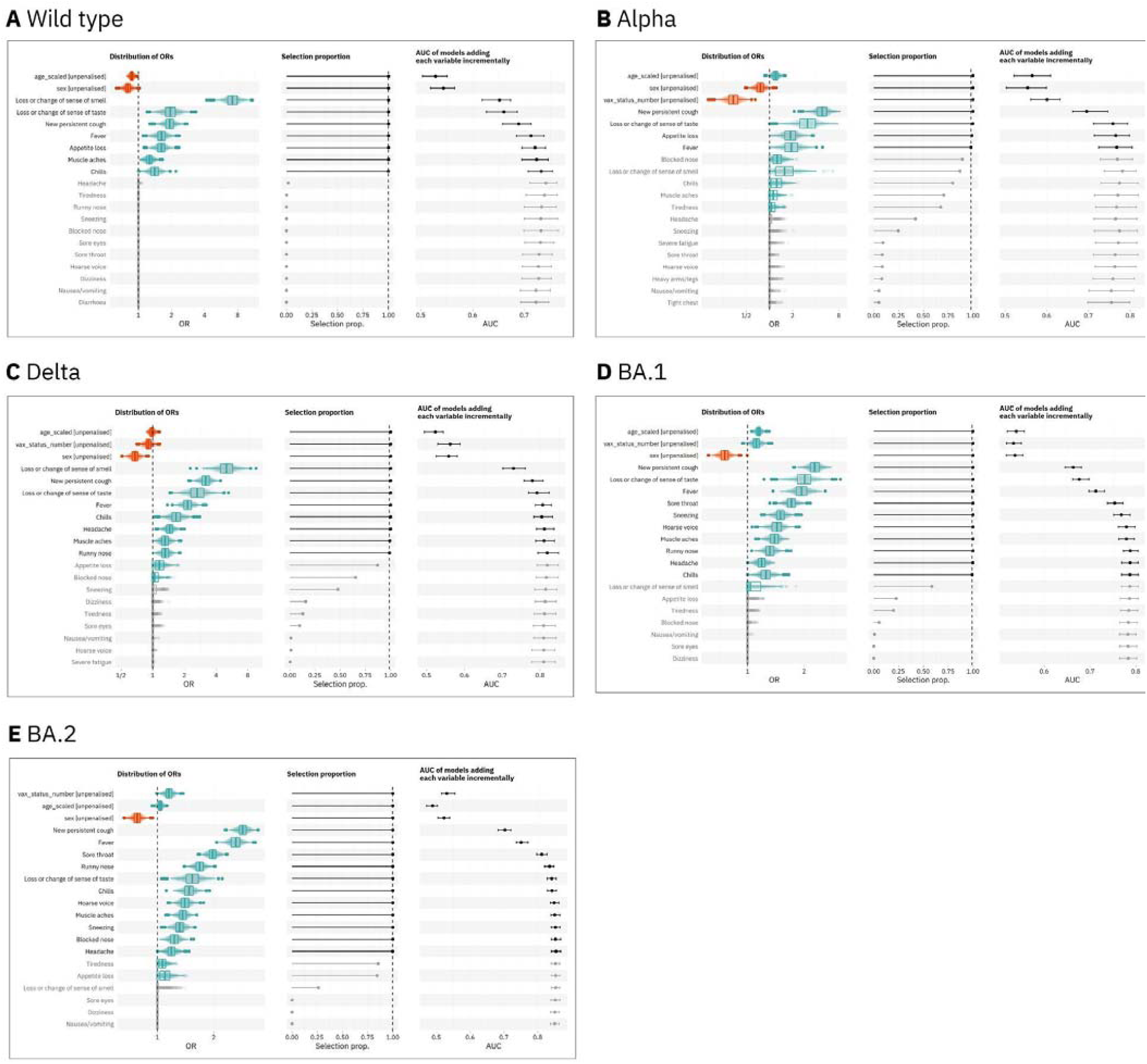
Results of LASSO stability selection with swab positive/negative as the binary outcome variable and each of 26 symptoms as predictors, in four phases of REACT-1 corresponding to periods of dominance of four SARS-CoV-2 variants in England. Age, sex and, where appropriate, vaccination status are forced into the models as unpenalised variables; regression coefficients for the symptoms are constrained to be positive. The median, 5th and 95th percentiles of AUCs are obtained from unpenalised models adding each symptom incrementally (from top to bottom) applied on un-seen holdout data. with regression coefficients for symptoms constrained to be non-negative.

**Figure S3.**
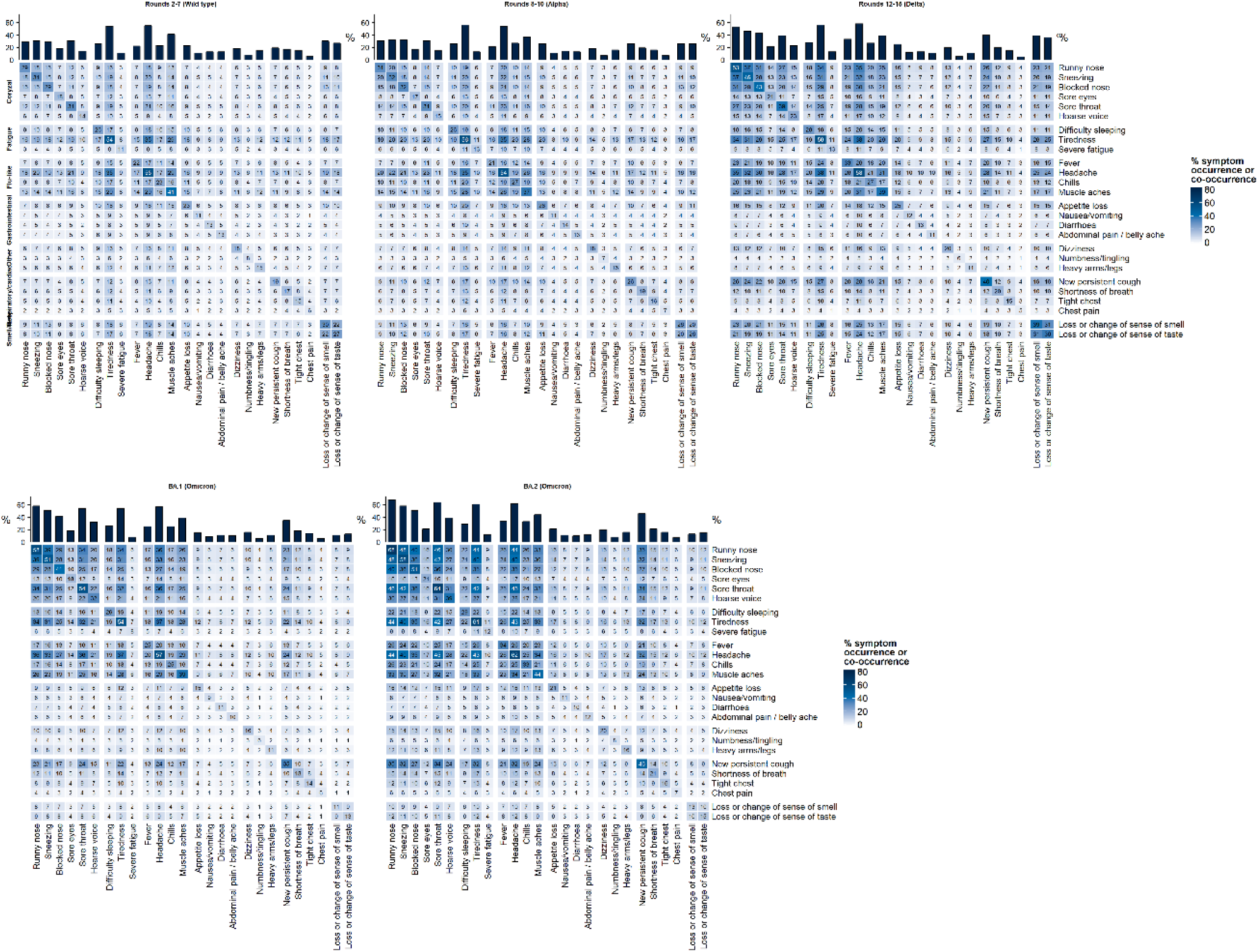
Heatmaps showing co-occurrence of symptoms across variants among swab-positive individuals. Denominators for percentages are the number of swab positive individuals with each variant.

**Figure S4.**
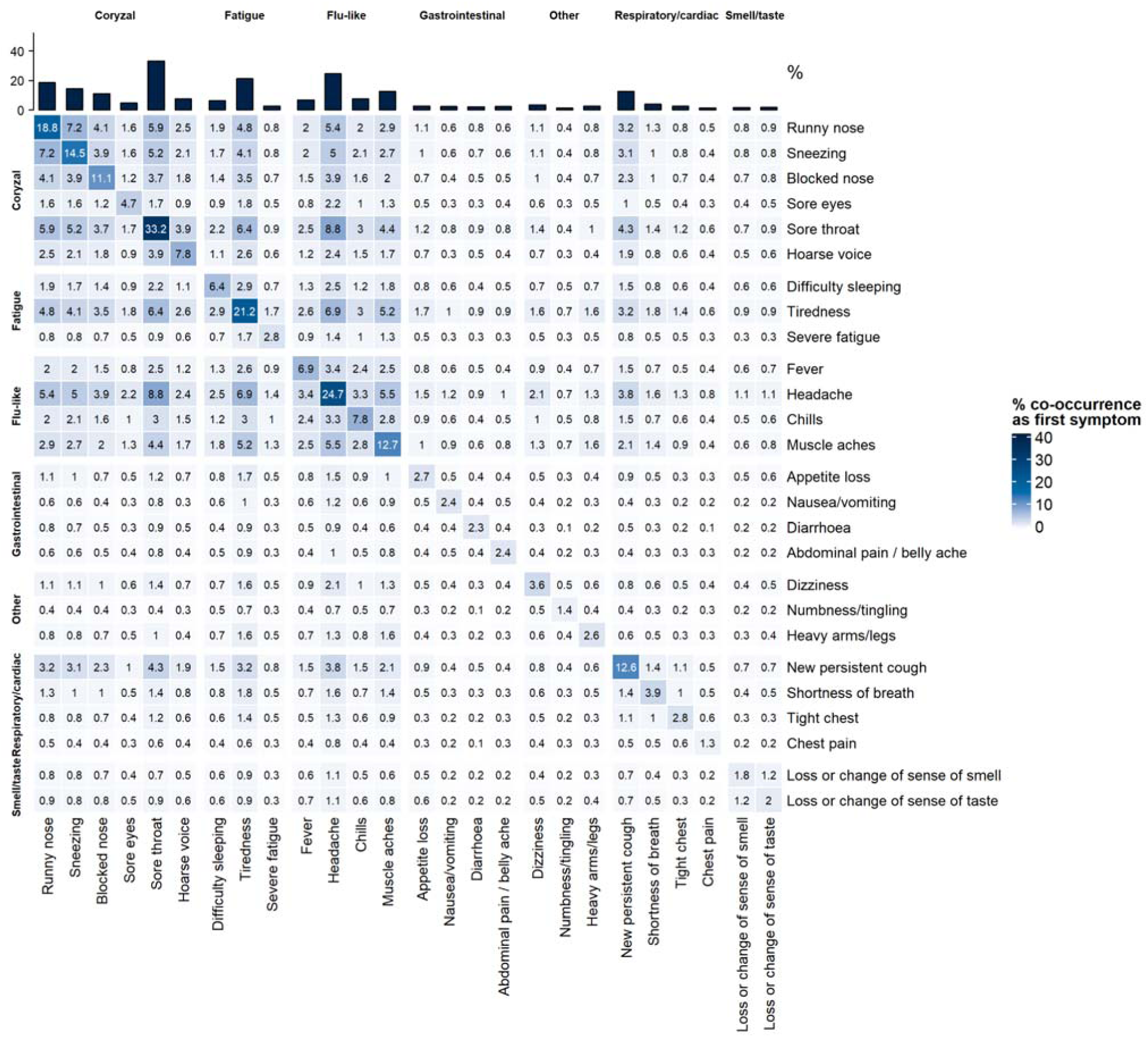
Heatmap showing co-occurence of first symptoms among swab positive individuals in rounds 17–19, when Omicron was dominant. The denominator for percentages is the number of swab positive individuals who reported experiencing one or more of 26 symptoms in the week prior to testing in rounds 17–19 (n=7,176). Sore throat, headache, and tiredness are the most common first symptoms, are are the most commonly co-occurring pairs of first symptoms.

**Figure S5.**
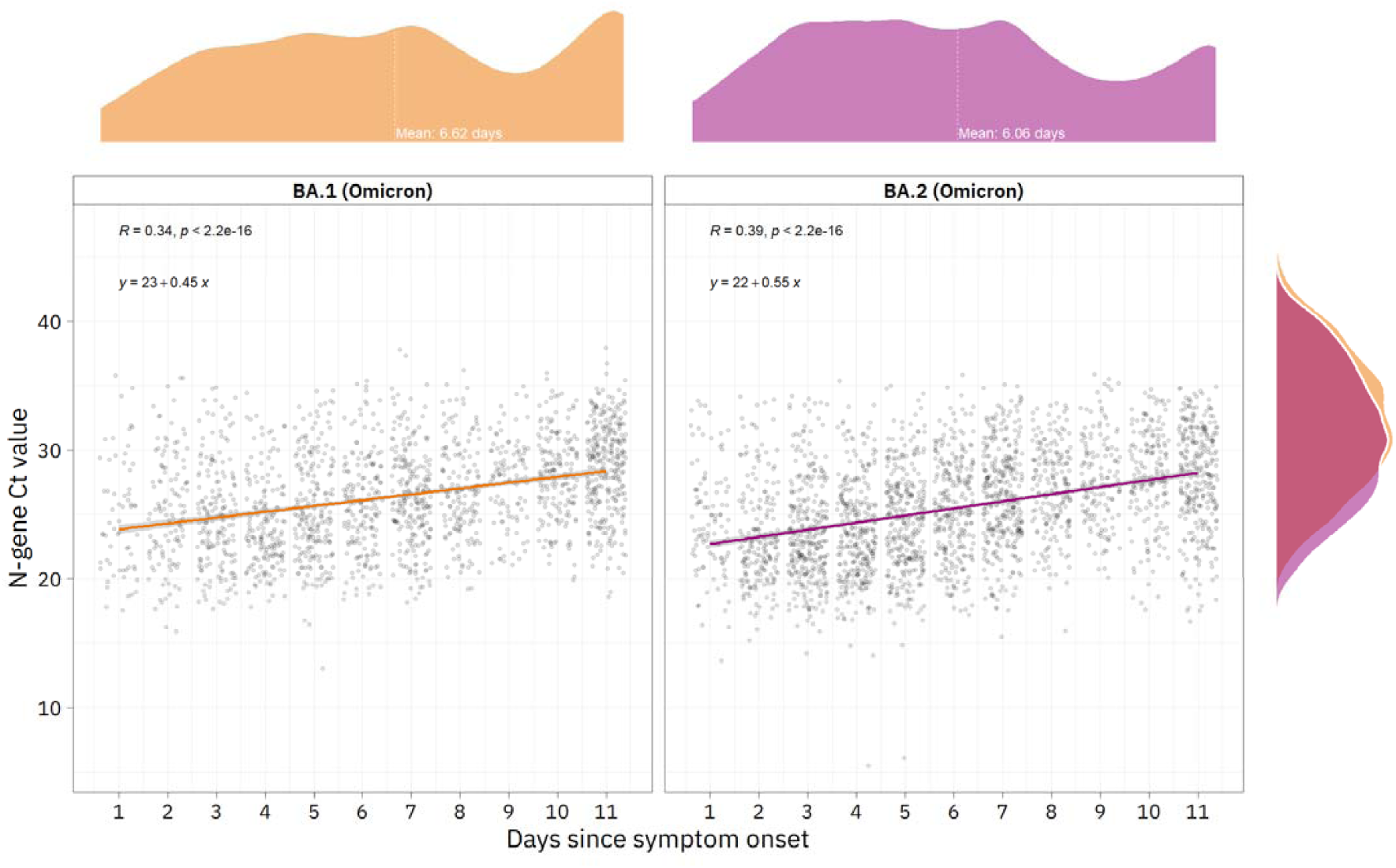
Scatter plot showing N-gene Ct values among symptomatic swab-positive individuals plotted against time (in days) since symptom onset, by BA.1/BA.2. Within each variant, a linear regression model is fitted with Ct value as the dependent variable and days since symptom onset (between 1 and 11 or more). In BA.2, there is a more even spread of time-since-symptom onset. In both variants there is a positive association between time since symptom onset and Ct value.

## Supplementary methods

### Assessing model performance in variable selection

To quantify the gain in predictive accuracy conferred by each selected symptom, un-penalised logistic models were successively re-fit on 80% subsamples of the holdout data, adding each symptom in order of decreasing selection proportion, and evaluated on the remainder of the holdout data (20%). Age, sex and vaccination status were forced in as predictors in all models. For each set of predictors, the procedure was repeated 100 times with different splits of the holdout data. As in the variable selection, regression coefficients were constrained to non-negativity in the re-fit models.

